# A Three-Item Functional Screen for Multimodal Prognostic Triage in Mild Cognitive Impairment: Benchmarking Against Entorhinal Tau PET and Plasma p-tau217

**DOI:** 10.64898/2026.06.01.26354584

**Authors:** Juliette Lafille, Frank Provenzano, Alzheimer’s Disease Neuroimaging Initiative

**Author notes:** **Correspondance**: Juliette Lafille, or Frank Provenzano. Data used in preparation of this article were obtained from the Alzheimer’s Disease Neuroimaging Initiative (ADNI) database (adni.loni.usc.edu). As such, the investigators within the ADNI contributed to the design and implementation of ADNI and/or provided data but did not participate in the analysis or writing of this report. A complete listing of ADNI investigators can be found at: http://adni.loni.usc.edu/wp-content/uploads/how_to_apply/ADNI_Acknowledgement_List.pdf.

## Abstract

**Importance:** Broadening access to biomarker-informed risk stratification in mild cognitive impairment (MCI) has become even more critical to early assessment in Alzheimer disease given recent developments in regulatory approvals of disease-modifying therapies and advancements of blood-based biomarkers. This requires accessible approaches that can be deployed at scale to better differentiate the disease biology from the clinical progression risk prediction. While entorhinal tau positron emission tomography (PET) can refine near-term prognostic assessment, the cost and logistic burden of imaging limit broad clinical use.

**Objective:** Evaluate whether a brief informant-reported screen derived from the Functional Activities Questionnaire (FAQ) could better stratify scalable biologically anchored prognostic information for 3-year progression from MCI to Alzheimer disease dementia. The primary study was designed around FAQ-derived screens performance relative to entorhinal tau PET standardized uptake value ratio (SUVR), plasma phosphorylated tau 217 (p-tau217) and Mini-Mental State Examination (MMSE) score. Secondary analyses evaluated the stable FAQ-derived screen selected for clinical risk separation, tau and amyloid PET biological context, additional plasma biomarkers, resource-use scenarios and sensitivity analyses around subgroups, calibration, decision-curve, survival, timing, early-progressor exclusions and endpoint-ascertainment IPW.

**Design, Setting, and Participants:** This retrospective secondary progression risk prediction study analyzed 350 Alzheimer’s Disease Neuroimaging Initiative (ADNI) participants with a baseline clinical diagnosis of MCI at the tau PET anchor visit. All studies were conducted in cohorts with 3-year progression status known. The first primary benchmarking included 157 participants (including 32 progressors) for FAQ with entorhinal tau PET SUVR comparisons and 153 participants (including 31 progressors) for FAQ, entorhinal tau PET SUVR and MMSE comparisons. The second primary benchmarking was derived from a smaller UPENN plasma p-tau217 subset of 66 participants (including 13 progressors).

**Exposures:** The FAQ-derived candidate screens were evaluated by leakage-controlled repeated nested cross-validation. The stable 3-item FAQ-derived screen selected was defined as any informant-reported difficulty in at least one of the three activities comprising finances/checkbook, shopping and games/hobbies (“Locked FAQ Trio”). The Locked FAQ Trio was compared against both biological and cognitive comparators: entorhinal tau PET SUVR, plasma p-tau217 and MMSE score. Amyloid PET status and Centiloid burden as well as plasma biomarkers paired per same-file plasma such as Aβ42/40 ratio, glial fibrillary acidic protein (GFAP), neurofilament light chain (NfL) and a directionally adjusted 4– marker plasma composite were used for biology or exploratory context and not for defining the clinical endpoint.

**Main Outcomes and Measures:** The primary binary endpoint was progression from baseline MCI at the tau PET anchor visit to Alzheimer disease dementia within 3 years. Model performance used the cross-validated area under the receiver operating characteristic curve (AUC), the difference in AUC (ΔAUC) was bootstrap 95% confidence intervals (CI) at the participant level with P values adjusted using the Benjamini-Hochberg (BH) procedure. Other measures included Brier scores, calibration summaries, survival discrimination and operating characteristics such as sensitivity, specificity, positive predictive value (PPV), negative predictive value (NPV) and screen-positivity prevalence, while decision-curve analyses and resource-use scenarios remained exploratory.

**Results:** A leakage-controlled nested cross-validation selection repeatedly identified a 3-item screen defined as any difficulty in at least one of the three following activities comprising finances/checkbook, shopping and games/hobbies (Locked FAQ Trio). In an independent 3-year progression benchmark analysis of base-covariate models, the Locked FAQ Trio showed higher numerical, directional but not statistically significant, discrimination than entorhinal tau PET among 157 participants including 32 progressors (AUC, 0.787 vs 0.780; ΔAUC, +0.007; 95% CI, −0.099 to 0.113; BH-adjusted P = 0.926) and was statistically significantly higher than MMSE score (AUC, 0.796 vs 0.637; ΔAUC, +0.159; 95% CI, 0.045 to 0.276; BH-adjusted P = 0.029). The Locked FAQ Trio was positive in 37.6% of participants and captured 27 of 32 progressors, showing sensitivity of 84.4%, specificity of 74.4%, PPV of 45.8%, and NPV of 94.9%. Progression within 3 years occurred in 45.8% of screen-positive participants versus 5.1% of screen-negative participants and the corresponding adjusted hazard ratio over full follow-up was 7.46. The screen was also associated with higher entorhinal tau burden and remained consistent across survival, timing-sensitive, amyloid and missingness analyses. A different 3-item FAQ-derived companion screen (“Companion FAQ Trio”) was evaluated for sensitivity, it was defined as any impairment in at least one of the three activities comprising forms/papers, shopping and remembering appointments/medications/holidays. The Companion FAQ Trio was positive in 54.1% participants and captured 96.9% of progressors, with 36.5% of screen-positive progressing to dementia versus 1.4% of screen-negative.

In a second primary benchmark analysis of a smaller matched plasma subset of 66 participants including 13 progressors, plasma p-tau217 showed the highest discrimination (AUC, 0.890) across all single predictors in a base-covariates model, compared with the Locked FAQ Trio (AUC, 0.749) and entorhinal tau PET SUVR (AUC, 0.798). A stratification study of the Locked FAQ Trio combined with p-tau217 showed separation of observed risk, differentiating lower and higher risk of progression per strata. Notably, none (0 of 31) of the participants in the lower risk cohort progressed and 64.3% (9 of 14) of participants in the higher risk cohort progressed. Nevertheless, 37.5% (3 of 8) of participants in the Locked FAQ Trio-negative/p-tau 217-high cohort progressed. This emphasizes that patients should not be excluded from further biomarker testing when clinical concern remains.

**Conclusion:** A brief 3-item stable FAQ-derived screen was identified as a compelling front-end additional layer to prognostic triage in MCI patients. This Locked FAQ Trio screen demonstrated a higher numerical discrimination than entorhinal tau PET SUVR in 3-year base-covariates prediction risk models. Plasma p-tau217 remained the strongest scalable predictor of progression to dementia in a smaller plasma subset. These findings reinforce that adding a brief functional screen to the staged prognosis assessment triage pathway can help prioritize and contextualize biomarker escalation, offering a scalable, deployable, and low burden solution to expand screening to a broader patient population.

**Key Points:** *Question:* Can a low-burden brief informant-reported functional questionnaire support staged prognostic triage, before biomarker escalation, for near-term progression risk from mild cognitive impairment to Alzheimer disease dementia?

*Findings:* In this progression risk prediction study of 350 individuals with mild cognitive impairment, a 3-item Functional Activities Questionnaire (FAQ) was identified as a stable early signal for progression risk using a leakage-controlled repeated nested cross-validation. The screen was defined as any impairment in at least one of the three activities comprising finances/checkbook, shopping and games/hobbies (“Locked FAQ Trio”). In an independent prognosis prediction study, the Locked FAQ Trio was numerically, but not statistically significantly, higher than entorhinal tau positron emission tomography (PET) standardized uptake value ratio (SUVR) and statistically significantly higher than Mini-Mental State Examination (MMSE) score. In a smaller plasma subset of 66 participants, plasma phosphorylated tau 217 (p-tau217) showed the highest discrimination and the Locked FAQ Trio combined with p-tau217 differentiated lower and higher risk of progression.

*Meaning:* An informant-reported brief 3-item functional questionnaire can help to inform and prioritize biomarker testing. A selected Locked FAQ Trio showed a higher numerical discrimination than specialized entorhinal tau PET biomarker and contextualized plasma p-tau217 biomarker. The suggested staged framework starts with Locked FAQ Trio screen triage, then plasma p-tau217 refinement before selective confirmation disease pathology with cerebrospinal fluid biomarkers or amyloid PET and/or tau PET for staging or prognostic prediction.

## Introduction

With recent developments in Alzheimer disease both in regulatory approvals of disease-modifying therapies targeting underlying pathology and in advancements of diagnostics, in particular blood-based biomarkers, the interest in broadening access to biomarker-informed risk stratification in Alzheimer disease has been increasing in recent years. The expert disease stratification criteria increasingly differentiates the disease biology status between clinical syndrome, staging, and prognosis [1], [2], [3]. This distinction is central to the present study which aims at stratifying the risk of near-term disease progression in individuals with baseline clinical mild cognitive impairment (MCI), rather than aiming at assessing disease biology.

MCI is heterogeneous and does not suffice to predict the progression risk to dementia as MCI patients experience a wide range of outcomes, with some progressing to dementia, others stabilizing and some improving [4], [5], [6]. To assess the near-term risk of progression in MCI patients, clinicians need to conduct more comprehensive evaluations beyond biology that include cognition, function, and broader clinical context (e.g.: age, education, vascular conditions). While core biomarkers like tau and amyloid positron emission tomography (PET) imaging or cerebrospinal fluid (CSF) have considerably improved prognosis evaluations, those are resource-intensive and difficult to scale. Additional staged lower burden tools that remain biology-anchored could further stratify clinical transition risk evaluations.

Alzheimer disease blood-based biomarkers are rapidly reshaping how clinicians evaluate symptomatic cognitive impairment with plasma phosphorylated tau 217 (p-tau217)-centered measures showing particularly strong accuracy in defining the disease pathology [7], [8], [9], [10], [11]. Recent regulatory developments support the use of those plasma biomarkers into clinical implementation but with some limitations. In 2025, the US Food and Drug Administration (FDA) cleared the Lumipulse G pTau217/β-Amyloid 1-42 Plasma Ratio, the first blood-based in vitro diagnostic device to aid identify amyloid pathology associated with Alzheimer disease in symptomatic patients in specialized-care settings. The scope remains narrow, it is assay-specific, limited to the clinic setting and does not support stand-alone screening, stand-alone diagnostic or treatment decisions based solely on the blood test [12]. Current guidance clearly distinguishes between blood-based biomarkers that support confirmatory use in selected settings and those that can help characterize the risk of progression [13], [14], [15].

High performing biomarkers like PET imaging or CSF collection can help determine near-term progression risk and staging in MCI patients, but their implementation as a PET-first or CSF-first diagnostic pathway is limited by accessibility, capacity and cost constraints. The recent appropriate use criteria is supportive of selective amyloid or tau PET scans in MCI patients when the results can change diagnostic confidence, staging, prognosis, management or trial eligibility, creating practical implementation challenges [16], [17]. To alleviate the operational burden, a staged triage pathway could identify MCI patients that would benefit from a prognosis assessment sooner. A practical pathway may begin with low burden clinical progression risk assessment, then scalable blood-based biomarker testing, followed by selective CSF and PET testing.

As the treatment paradigm continues to evolve, progression risk stratification and evaluation of early symptomatic patients become even more consequential. The FDA recently approved disease-modifying treatments lecanemab and donanemab in MCI or mild dementia settings. On one hand, low burden prognostic assessment could serve as a scalable front-end triage in MCI patients who would have not tested for progression risk otherwise, thus detecting additional patients at higher risk of near-term decline who may test plasma, CSF or PET biomarkers earlier, as well as engage in treatment-related counseling. On the other hand, the availability of disease-modifying therapies may itself incentivize patients and clinicians to seek earlier prognostic and biomarker assessment as those results can inform whether to pursue more resource-intensive testing and treatment counseling. The current prescribing labels of lecanemab and donanemab require confirmation of the presence of amyloid-β (Aβ) pathology prior to initiating treatment, staging, magnetic resonance imaging (MRI) safety assessment and individualized risk discussions [18], [19], [20], [21]. While prognostic stratification does not determine treatment eligibility itself, it may inform urgency and prioritization of downstream evaluation.

The Functional Activities Questionnaire (FAQ) is a natural candidate for low burden front-end triage and contextualization. FAQ captures informant-reported changes in real-world function, which is observable, low-cost and deployable at scale without the need for laboratory or assay. More importantly, FAQ is designed around instrumental activities of daily living (IADLs), a clinically meaningful domain where subtle losses often become visible near the MCI to dementia threshold. Given this, the FAQ is not viewed as a biomarker or treatment eligibility substitute, but rather as an additional pre-biomarker clinical layer, grounded in real-world functional vulnerability [22], [23], [24], [25], [26], [27], [28], [29].

Using carefully characterized ADNI data [30], [31], we tested whether a small set of FAQ-derived items could support scalable biologically anchored progression risk triage for 3-year progression from MCI to Alzheimer disease dementia. In the primary progression risk prediction study, we used a leakage-controlled nested cross-validation model to evaluate brief FAQ-derived screens against entorhinal tau PET (standardized uptake value ratio) SUVR, a specialized imaging benchmark for tau staging and prognosis, and Mini-Mental State Examination (MMSE) score, a brief cognitive comparator [16], [17], [32]. This procedure repeatedly selected a stable stand-alone 3-item FAQ-derived screen defined as any impairment in at least 1 of the 3 activities comprising finances/checkbook, shopping and games/hobbies (“Locked FAQ Trio”). We then compared this stable Locked FAQ Trio as the predictor in independent progression risk prediction analyses against entorhinal tau PET SUVR (Aim 1), MMSE and plasma p-tau217 (Aim 2) in the source-matched plasma subset while retaining entorhinal tau PET SUVR as the imaging benchmark on the same denominator [7], [8], [9], [10], [11], [13], [14], [15]. In secondary analyses, we also evaluated the Locked FAQ Trio for clinical risk separation, sensitivity analyses, biological context and resource-use scenario, using amyloid PET status and Centiloid burden as well as plasma biomarkers paired per same-file such as Aβ42/40 ratio, glial fibrillary acidic protein (GFAP), neurofilament light chain (NfL) and a directionally adjusted 4– marker plasma composite. The aim was to test whether a brief informant-reported functional signal could support front-end clinical triage of near-term progression risk from MCI to dementia and guide more urgent selective downstream biomarker evaluation or specialist referrals for a broader MCI patient population.

## Methods

### Study Design and Data Source

Data used in the preparation of this article were obtained from the Alzheimer’s Disease Neuroimaging Initiative (ADNI) database (adni.loni.usc.edu). The original goal of ADNI was to test whether serial magnetic resonance imaging (MRI), positron emission tomography (PET), other biological markers and clinical and neuropsychological assessment can be combined to measure the progression of MCI and early Alzheimer’s disease (AD). The current goals include validating biomarkers for clinical trials, improving the generalizability of ADNI data by increasing diversity in the participant cohort and to provide data concerning the diagnosis and progression of Alzheimer’s disease to the scientific community. For up-to-date information, see adni.loni.usc.edu.

This retrospective secondary study of ADNI dataset evaluated prognostic prediction and comparator benchmarking for near-term progression from MCI to Alzheimer disease dementia [30], [31]. The study cohorts included participants with clinical MCI at baseline and a tau PET scan visit anchor date. The analytic baseline was anchored to each participant’s first available tau PET scan, the main imaging benchmark in the study. The binary endpoint was defined as the progression status from MCI to Alzheimer disease dementia within 3 years from the tau PET anchor. Progression status was also obtained from the ADNI dataset. The study was designed as a prognostic prediction model with FAQ-derived screens as the primary clinical predictor and entorhinal tau PET SUVR (Aim 1), plasma p-tau217 (Aim 2) and MMSE score as comparators in discrimination analyses. Multimodal discriminations and progression risk stratifications were evaluated in the context of possible front-end triage to further downstream biomarker testing and not as a substitute to diagnosis, staging or treatment eligibility. To avoid optimistic performance estimates, the FAQ-derived screen discovery procedure was evaluated independently from the performance evaluation and modeled with a leakage-controlled nested cross-validation framework. Secondary, exploratory and contextual studies included Amyloid PET burden and additional plasma biomarkers paired per same-file such as Aβ42/40 ratio, GFAP, NfL and a directionally adjusted 4– marker plasma composite. Sample sizes varied according to availability of the relevant outcomes and predictors and denominators are non-interchangeable.

Reporting followed TRIPOD+AI as the primary prediction model framework, we also used STROBE for observational context, PROBAST/PROBAST+AI for internal risk of bias and self-auditing and STARD for descriptive clinical interpretation [33], [34], [35], [36].

### Analytic Hierarchy

The prediction model study was designed around two primary benchmarking analyses and several exploratory secondary studies such as biological context, sensitivity, resource-use and source audit analyses. The primary Aim 1 first identified a brief FAQ-derived screen in a leakage-controlled repeated nested cross validation and then evaluated independently its held-out prediction performance against the tau-anchored and endpoint-known entorhinal tau PET SUVR. The benchmarking Aim 1 analysis reflects the expected performance of the selected screen candidate rather than the apparent performance of a rule selected and evaluated in the full dataset.

The primary Aim 2 evaluated the Locked FAQ Trio against UPENN plasma p-tau217 in a smaller plasma available complete-case subset with the entorhinal tau PET SUVR benchmark also evaluated on the same denominator as another comparator. The Locked FAQ Trio was retained from the Aim 1 analysis and we did not reopen the FAQ screen discovery in the Aim 2 analysis given the limited size of the subset.

Secondary analyses evaluated fixed Locked FAQ Trio performance, operating characteristics, calibration, survival discrimination, timing robustness, early-progressor exclusions and decision curves. Biological context analyses examined the alignment between the FAQ screens and entorhinal tau PET and amyloid PET biology. Source audit and diagnostic resource-use analyses remained exploratory. MMSE score served as a clinical benchmark comparator within the primary Aim 1 given data present within the same 350 MCI participants cohort.

### Cohort Construction

We identified each participant’s earliest available tau PET scan visit and used it as the anchor date. We linked this tau PET anchor visit to the closest diagnosis visit within 365 days and preferred a diagnosis on or before the tau PET date when available. We only retained participants with MCI diagnosis at the anchor tau PET visit. We then paired each anchor tau PET scan visit with the closest FAQ visit within 180 days. This cohort construction included 350 participants with a tau anchored date, MCI diagnosis at baseline and FAQ data available.

Within this cohort of 350 participants, we derived the binary 3-year analytic cohort from longitudinal diagnosis follow-up. We counted participants as endpoint-known either if they progressed to dementia diagnosis within 3 years or if they completed at least 3 years of follow-up without dementia. We counted the participants who had shorter follow-up and no dementia diagnosis as endpoint-unknown for the primary binary analyses. Following this cohort construction process, we counted 163 tau-anchored MCI participants with 3-year status endpoint-known, of which 34 had progressed to dementia within 3 years and 129 had not progressed. This 163 participants subgroup was used for denominator accounting and reflects endpoint ascertainment rather than predictor missingness. Each primary benchmark analysis used its own denominator as each cohort was adjusted according to respective comparators data availability.

The primary Aim 1 analysis benchmarking FAQ to entorhinal tau PET SUVR counted 157 of those 163 participants, including 32 of those 34 progressors, and the benchmarking of FAQ to MMSE score evaluated 153 of those 163 participants, including 31 of those 34 progressors. In a smaller UPENN plasma p-tau217 subset, the primary benchmarking analysis Aim 2 counted a model-specific subset of 66 participants including 13 progressors.

### Outcome

The primary endpoint was the progression from MCI to Alzheimer disease dementia within 3 years of the tau PET anchor visit, analyzed as a binary outcome among participants with known 3-year progression status. While participants without sufficient follow-up to determine 3-year progression status were excluded from the binary analyses, they remained eligible for full follow-up time-to-event analyses. For survival analyses, the follow-up time was measured in days from the tau PET anchor visit to the visit date of conversion progression for participants who progressed and to the last available follow-up for those who did not.

### Functional Activities Questionnaire Predictors and Screen Construction

We harmonized the 10 FAQ items answers to a 0-to-3 scale and derived the FAQ total score when all 10 items contributed [22]. We defined GE1 (Greater or Equal to 1) as any difficulty on any item, corresponding to item score ≥1. We defined GE2 (Greater or Equal to 2) as a stricter threshold corresponding to an item score ≥2 on the scale of 0-to-3. We generated candidate screens from the 10 single FAQ items, any 2, 3 or 4 combinations among all 10 items and we also considered the FAQ total sum scores with thresholds of ≥6 and ≥9. The full candidate screens library evaluated 774 screens or 387 screens at the GE1 threshold and 387 screens at the GE2 threshold. For each GE threshold, the candidate screens comprised 10 single-item screens, 2 total-score sum, 45 any-of-2 rules, 120 any-of-3 rules and 210 any-of-4 rules. FAQ item domains and harmonized response scoring are shown in eTable 2.

The leakage-controlled screen selection procedure repeatedly prioritized a GE1 3-item screen comprising of finances/checkbook, shopping and games/hobbies (Locked FAQ Trio) and we carried this rule forward. The Locked FAQ Trio classified a participant as screen positive when the participant had any difficulty on at least one of those three items. We also examined a companion GE1 screen as well as FAQ total sum at the ≥6 threshold to support clinical interpretation of sensitivity, specificity, predictive values, and testing-escalation tradeoffs. The companion GE1 screen was defined as any difficulty in at least 1 of the 3 activities comprising forms/papers, shopping and remembering appointments/medications/holidays (“Companion FAQ Trio”).

### Imaging, Cognitive, Plasma and Covariate Predictors

The present study reflects a multimodal analysis comparing function, cognition, tau PET, and plasma biomarkers in one framework. We used two biological comparators in the primary study, entorhinal cortex tau PET SUVR (Aim 1) as imaging benchmark [16], [17] and UPENN plasma p-tau217 (Aim 2) as scalable plasma benchmark [7], [8], [9], [10], [11], [13], [14], [15]. In addition to biological comparators, we used MMSE score as a brief cognitive comparator [32]. We also evaluated paired source-matched plasma Aβ42/40, GFAP, and NfL in secondary or exploratory models only. Amyloid PET status and Centiloid burden contextualized biology and were not evaluated as primary comparators for the FAQ-derived screen selection procedure [16], [37]. Base covariates included age at tau PET, sex, education and apolipoprotein E ε4 (APOE ε4).

### Primary Benchmarking Analysis Aim 1: Leakage-Controlled FAQ Screen Selection

The primary benchmarking analysis Aim 1 used four different blocks for comparisons. Block A compared the base covariates plus the selected FAQ screen with the base covariates plus entorhinal tau PET SUVR. Block B compared the base covariates plus the selected FAQ screen with the base covariates plus the MMSE score. Block C evaluated the incremental value of the selected FAQ screen beyond the base covariates plus entorhinal tau PET SUVR. Similarly, Block D evaluated the incremental value of the selected FAQ screen beyond the base covariates plus entorhinal tau PET SUVR plus MMSE score. The FAQ screen was selected within the training data of each outer cross-validation split in each block.

We used repeated nested cross-validation to control leakage during screen selection. Each outer training fold contained an inner cross-validation loop that selected one FAQ screen from the candidate library by mean inner-loop area under the receiver operating characteristic curve (AUC), with a prespecified parsimony tie rule. The outer loop fit the selected model in the outer training fold and generated held-out predictions for the outer test fold. We repeated the procedure across 5 outer folds, 5 inner folds, and 20 repeats. We averaged repeated held-out predictions within participant and used the participant-level predictions for AUC, Brier score, calibration, decision-curve summaries, and paired bootstrap comparisons.

### Locked FAQ Trio Sensitivity Analyses

We analyzed the Locked FAQ Trio as an internal post-selection sensitivity analysis. We did not treat this analysis as external validation. We fit the same block-specific model structures with the Locked FAQ Trio by replacing the selected screen. We calculated the participant level cross-validated AUCs, Brier scores, paired ΔAUCs, bootstrap 95% confidence intervals, and Benjamini-Hochberg (BH) adjusted P values across the four primary GE1 blocks (A-D) ΔAUC tests [38]. We also described the operating characteristics for the Locked FAQ Trio as well as for the Companion FAQ Trio and the FAQ total sum at ≥6 threshold, including sensitivity, specificity, positive predictive value (PPV), negative predictive value (NPV) and escalations per converter captured. We used Kaplan-Meier curves and Cox proportional hazards models to examine risk separation over full follow-up [39].

### Primary Benchmarking Analysis Aim 2: Plasma p-tau217 Benchmarking

We evaluated the plasma models based on the UPENN p-tau217 complete-case smaller subset on a common denominator of 66 participants, including 13 progressors. We matched the plasma sampling to the tau PET anchor within 90 days. We fit the fixed logistic models for base covariates alone, base plus Locked FAQ Trio, base plus entorhinal tau PET SUVR, base plus p-tau217, and combined FAQ, tau PET, and p-tau217 models. We used repeated stratified cross-validation, training-fold imputation and standardization, participant-level averaged held-out predictions, AUCs, Brier scores, and paired bootstrap ΔAUCs. We defined p-tau217-high as the internal top tertile of the UPENN p-tau217 z score in the final binary complete-case subset and p-tau217-not high as the internal bottom two tertiles. We cross-classified participants by Locked FAQ Trio status and p-tau217-high status and calculated the binomial confidence intervals for the 3-year progression proportion within each stratum. We did not treat the tertile threshold as a clinical cutoff.

### Secondary and Exploratory Analyses

Amyloid PET was analyzed only to contextualize the biology. We matched amyloid PET to the tau anchor visit date by participant and the nearest scan date. We used amyloid status when available and Centiloid burden when analyses required a continuous amyloid measure [16], [37]. These analyses evaluated whether FAQ and p-tau217 strata aligned with amyloid burden and we did not evaluate FAQ as a substitute for amyloid confirmation.

We audited p-tau181 and p-tau231 availability before modeling. We searched candidate plasma sources and candidate columns, matched candidate measures to the tau PET anchor across prespecified windows and required a model-ready denominator with both events and nonevents before estimating predictive models. When the selected p-tau181/p-tau217 common denominator lacked events, we did not estimate p-tau181 predictive models, correlations, or risk strata.

We evaluated robustness through endpoint-known versus endpoint-unknown profiling, inverse-probability weighted sensitivity analyses, timing-window analyses, missing-as-unimpaired FAQ sensitivity, no-APOE ε4 sensitivity, and early-progressor exclusions at 90 and 180 days. We treated inverse-probability weighting (IPW) as a diagnostic sensitivity analysis and decision-curve analyses and resource-use scenarios as exploratory translations of clinical testing volume and not as cost-effectiveness or treatment-access models [40], [41].

## Results

### Analytic Contexts, Cohort Composition and Timing Alignment

This study is presented across several analyses with their specific denominator which should not be transferred across other tables. Primary Aim 1 was analyzed in endpoint-known of 3-year status progression anchored at tau PET visit cohorts for comparisons between FAQ-derived screens with entorhinal tau PET SUVR and MMSE scores. Primary Aim 2 used a similar methodology but derived from a smaller UPENN plasma p-tau217 cohort. Evaluation of entorhinal tau and amyloid burden versus tau and amyloid biology used data where available. An audit of p-tau181/p-tau231 were used for source-availability audit as matched endpoint-known cohorts lacked adequate events for modeling. Diagnostic resource-use scenarios rescaled the operating characteristics from Aim 1 and Aim 2 denominators into per 1,000 testing volume scale. Cohort characteristics, endpoint-known accounting, and model-specific denominators are summarized in Table 1.

**Table 1.** Baseline Characteristics and Analytic Denominators across Primary Benchmarking Cohorts.

| Characteristic | Tau-anchored cohort (n=350) | Primary Benchmarking Analysis 1 A/C (n=157) | Primary Benchmarking Analysis 1 B/D (n=153) | Primary Benchmarking Analysis 2 p-tau217 (n=66) |
| --- | --- | --- | --- | --- |
| 3-y status known | 163 (46.6%) | 157 (100.0%) | 153 (100.0%) | 66 (100.0%) |
| 3-y progressions among endpoint-known | 34/163 (20.9%) | 32/157 (20.4%) | 31/153 (20.3%) | 13/66 (19.7%) |
| Age, mean (SD), y | 73.9 (7.9) | 73.7 (7.6) | 73.7 (7.6) | 72.8 (7.6) |
| Women, No. (%) | 149 (42.6%) | 56 (35.7%) | 55 (35.9%) | 24 (36.4%) |
| Education, mean (SD), y | 16.2 (2.6) | 16.4 (2.6) | 16.4 (2.6) | 16.1 (2.3) |
| APOE ε4 carrier, No. (%) | 133 (40.9%) | 65 (41.4%) | 62 (40.5%) | 26 (39.4%) |
| MMSE, mean (SD) | 27.8 (2.0); n=332 | 27.8 (2.0); n=153 | 27.8 (2.0) | 27.6 (2.2); n=65 |
| Entorhinal tau PET SUVR, mean (SD) | 1.314 (0.306); n=347 | 1.300 (0.262) | 1.299 (0.262) | 1.309 (0.272) |
| FAQ total score, mean (SD) | 3.1 (4.2); n=341 | 3.1 (4.3) | 3.1 (4.2) | 3.3 (4.6) |
| Locked FAQ Trio positive, No. (%) | 128 (37.5%); n=341 | 59 (37.6%) | 57 (37.3%) | 27 (40.9%) |
| plasma p-tau217 raw, median (range) | — | — | — | 0.166 (0.043-1.434) |

**Note:** Values are represented as Numbers (%) unless otherwise indicated. FAQ total score is the strict sum of 10 harmonized item scores, ranging from 0 to 30. UPENN plasma p-tau217 raw values are summarized only for the plasma model-specific subset and units follow the UPENN data dictionary. Analytic denominators are context-specific and should not be pooled.
**Abbreviations:** APOE ε4, apolipoprotein E ε4; FAQ, Functional Activities Questionnaire; MMSE, Mini-Mental State Examination; PET, positron emission tomography; p-tau217, phosphorylated tau 217; SD, standard deviation; SUVR, standardized uptake value ratio; UPENN, University of Pennsylvania.

This prognostic predictive study compared brief FAQ-derived screens with entorhinal Tau PET SUVR, plasma p-tau217 and MMSE scores predictors with participants anchored at tau PET visit. FAQ screens were compared to entorhinal Tau PET SUVR and MMSE scores in a cohort of 350 participants who harbored MCI at baseline and had a tau PET visit date anchor. Among them, 163 participants had an endpoint-known 3-year status, of which 20.9% (34/163) had progressed to Alzheimer disease dementia within 3 years. The endpoint was defined as the binary progression status at 3-year from the tau PET anchor visit. In primary Aim 1 the paired analyses with entorhinal Tau PET SUVR as a predictor (Blocks A/C) comprised 157 participants including 32 progressors and the paired analyses with MMSE scores as a predictor (Blocks B/D) comprised 153 individuals including 31 progressors. In primary Aim 2 the plasma benchmarking comprised 66 participants including 13 progressors, those derived from a smaller UPENN plasma subset with MCI at baseline, endpoint-known status, plasma p-tau measured within 90 days of the tau PET anchor visit as well as FAQ scores for finances/checkbook, shopping and games/hobbies (Locked FAQ Trio). Given the plasma subset was small, we used the fixed Locked FAQ Trio that emerged from the primary Aim 1 study and did not reopen the screen discovery in primary Aim 2. Baseline characteristics and model-specific denominators are shown in Table 1 and descriptive baseline contrasts by 3-year progression status are shown in eFigure 5.

Endpoint-know status at 3-year drove the denominator loss and data completeness remained high among predictors, with entorhinal tau PET SUVR available in 347/350, all 10 FAQ items in 341/350 and MMSE scores in 332/350, a missingness audit is available in eTable 1. Participants were comparable across variables, albeit a higher proportion of men 201/350 versus women 149/350 were included in the analyses.

FAQ assessments were obtained close to the tau PET anchor visit with median interval of 15.0 days with 64.6% (226/350) of participants assessed within 30 days and 92.6% (324/350) within 90 days. MMSE assessments were often obtained earlier and further from the anchor with a median interval of –47.5 days with 28.3% (94/332) assessed within 30 days and 80.1% (266/332) within 90 days. These distributions showed that FAQ assessments were generally acquired closer to the tau anchor than MMSE and support the rationale for further prespecified timing sensitivity analyses reported below. Timing distributions relative to the tau PET anchor date are shown in eFigure 9 and timing window sensitivity analyses are reported in eTable 12.

### Emergence of a Stable Brief FAQ-Derived Screen and Primary 3-Year Discrimination

A 3-item screen emerged from a nested cross validation repeated outer-loop selection at the primary GE1 threshold. The screen consisted of any reported difficulty in at least 1 of those 3 activities finances/checkbook, shopping and games/hobbies (Locked FAQ Trio) was selected the most frequently in Block A and independently again in Block B. These findings were supported by individual items selections with 92, 65 and 57 selections out of 100 outer-loop resamples in Block A and 95, 70 and 61 in Block B for finances/checkbook, shopping and games/hobbies, respectively. This Locked FAQ Trio was the most frequently selected in both binary and survival analyses, with binary stable shares of 0.31 in Block A and 0.32 in Block B and survival stable shares of 0.40 in Block A and 0.40 in Block B. The trio also ranked the highest by AUC in both blocks. The 3 activities selected are centered on planning, sequencing and judgement, those are instrumental to real world independence which may explain their sensitivity to the transition from MCI to dementia. Similarly, the close contender screens were coherent with the Locked FAQ Trio functions as they included a simpler 2-item screen of finances/checkbook and games/hobbies and a 4-item screen of finances/checkbook, shopping, games/hobbies and meal preparation. The screen selection stability is summarized in eTable 3 and item selection frequencies are visualized in eFigure 1.

**Figure 1.**
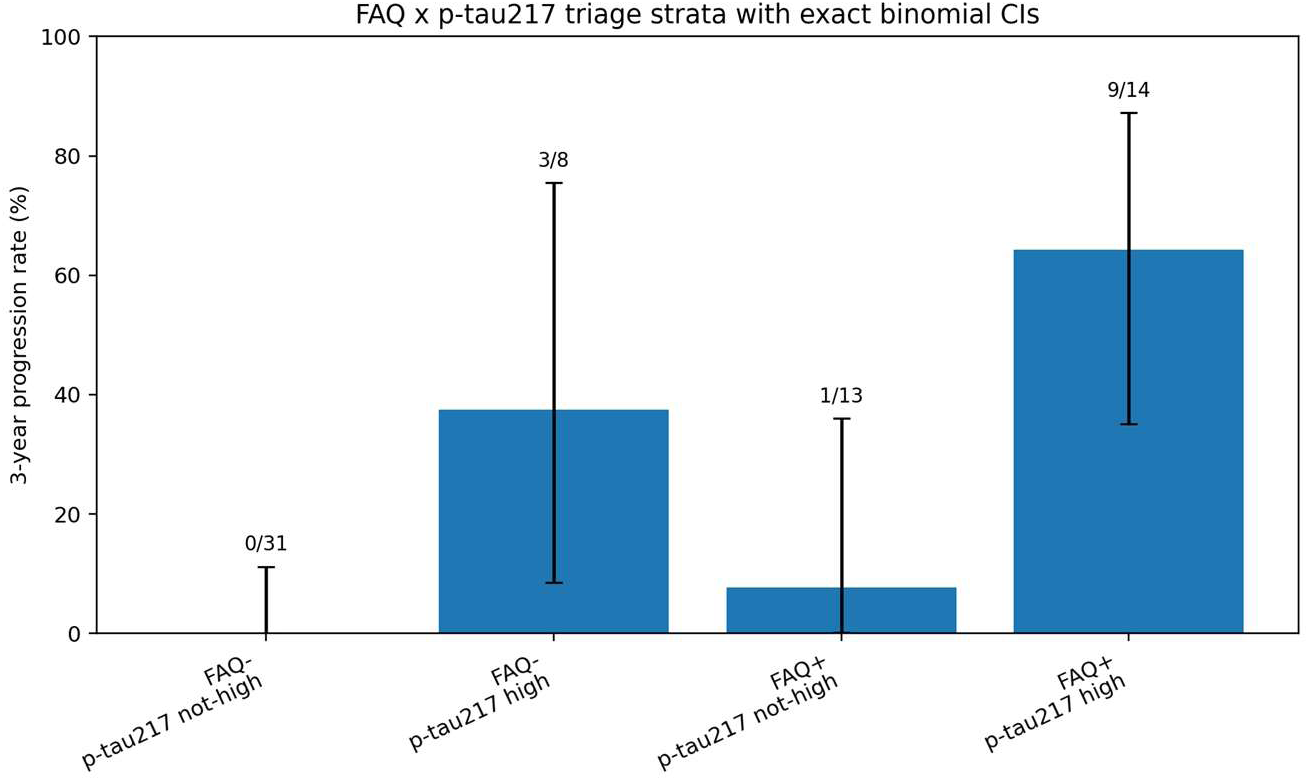
Locked FAQ Trio-by-p-tau217 Progression-Risk Strata in the UPENN Plasma Subset. **Note:** p-tau217-high was internally defined as the top tertile and p-tau217-not high as the bottom two tertiles of UPENN plasma p-tau217 in the final binary plasma subset. Error bars show exact binomial 95% confidence intervals. Sparse denominators preclude clinical cutoff claims. Some FAQ-negative/p-tau217-high participants still progressed, suggesting that FAQ status should not serve as a sole gatekeeper that prevents biomarker testing. **Abbreviations:** CI, confidence interval; FAQ, Functional Activities Questionnaire; p-tau217, phosphorylated tau 217; UPENN, University of Pennsylvania.

At the stricter difficulty threshold GE2, a similar screen selection model did not converge towards a compact multi-item combination screen, but instead, favored the summed-score FAQ total score of ≥6. Given that the GE2 threshold discrimination appeared overall less favorable than that of GE1, we interpreted GE2 analyses as secondary severity sensitivity analyses. GE2 screen selection behavior and full GE1/GE2 discrimination are shown in eTable 3 and eTable 4.

The IPW diagnostics for observed endpoint status appeared stable with no severe weight instability, the median weight reached 1.85, the 95th percentile weight reached 2.60 and the maximum weight reached 3.66 with the effective sample size reaching 147.8 among the 153 weighted endpoint-known participants.

### Primary Aim 1: Leakage-Controlled FAQ Screen Selection

Using a leakage-controlled nested cross-validation prediction framework, the GE1 selected FAQ screens showed a numerically higher discrimination for 3-year progression than the entorhinal tau PET SUVR comparator. In Block A, Base + selected FAQ screen reached an AUC of 0.787 versus 0.780 for Base + entorhinal tau PET SUVR (AUC, 0.787 versus 0.780; ΔAUC, +0.007; 95% CI –0.099 to +0.113; BH-adjusted P = 0.926). In Block B, the model also favored the GE1 selected FAQ screens with a statistically significant discrimination over MMSE scores (AUC, 0.796 versus 0.637; ΔAUC, +0.159; 95% CI +0.045 to +0.276; BH-adjusted P = 0.029). In incremental prognostic value analyses, again selected FAQ screens were favored, adding directionally positive incremental discrimination beyond entorhinal tau PET SUVR (ΔAUC of +0.059) beyond tau PET alone and beyond tau PET and MMSE (ΔAUC of +0.051). Primary GE1 selected screen discrimination results are summarized in Table 2 and full GE1/GE2 results are in eTable 4.

**Table 2.**
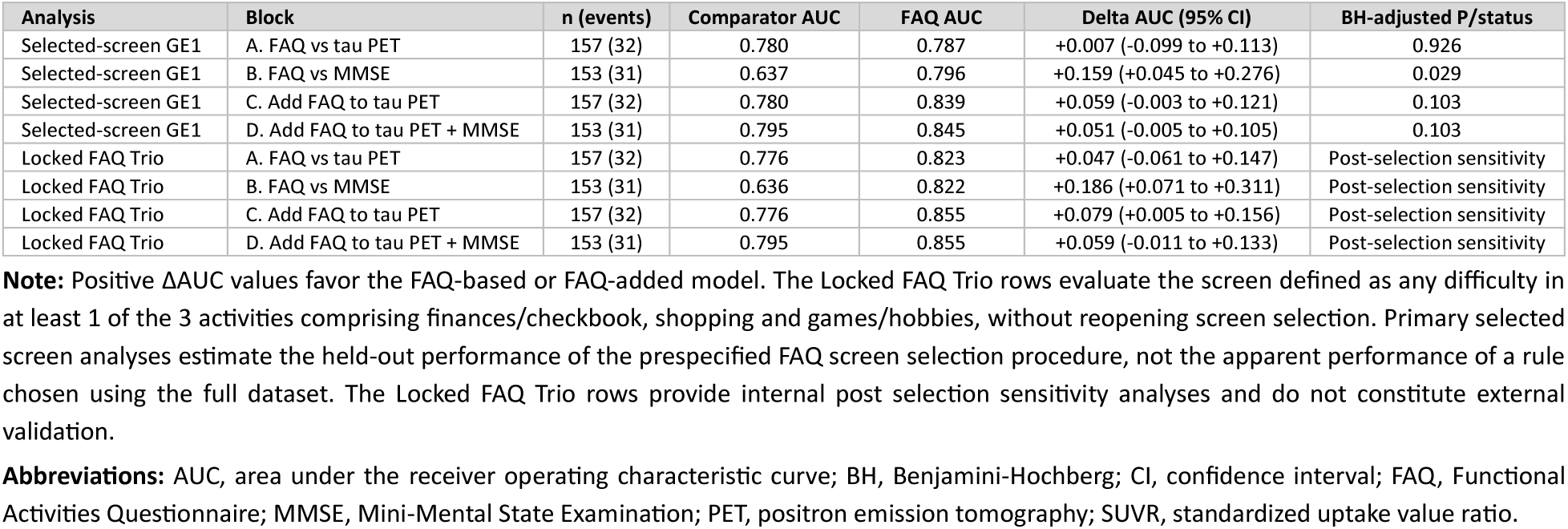
Primary Benchmarking Analysis Aim 1 Nested Cross-Validated Discrimination and Post Selection Fixed Locked FAQ Trio Sensitivity.

| Analysis | Block | n (events) | Comparator AUC | FAQ AUC | Delta AUC (95% CI) | BH-adjusted P/status |
| --- | --- | --- | --- | --- | --- | --- |
| Selected-screen GE1 | A. FAQ vs tau PET | 157 (32) | 0.780 | 0.787 | +0.007 (-0.099 to +0.113) | 0.926 |
| Selected-screen GE1 | B. FAQ vs MMSE | 153 (31) | 0.637 | 0.796 | +0.159 (+0.045 to +0.276) | 0.029 |
| Selected-screen GE1 | C. Add FAQ to tau PET | 157 (32) | 0.780 | 0.839 | +0.059 (-0.003 to +0.121) | 0.103 |
| Selected-screen GE1 | D. Add FAQ to tau PET + MMSE | 153 (31) | 0.795 | 0.845 | +0.051 (-0.005 to +0.105) | 0.103 |
| Locked FAQ Trio | A. FAQ vs tau PET | 157 (32) | 0.776 | 0.823 | +0.047 (-0.061 to +0.147) | Post-selection sensitivity |
| Locked FAQ Trio | B. FAQ vs MMSE | 153 (31) | 0.636 | 0.822 | +0.186 (+0.071 to +0.311) | Post-selection sensitivity |
| Locked FAQ Trio | C. Add FAQ to tau PET | 157 (32) | 0.776 | 0.855 | +0.079 (+0.005 to +0.156) | Post-selection sensitivity |
| Locked FAQ Trio | D. Add FAQ to tau PET + MMSE | 153 (31) | 0.795 | 0.855 | +0.059 (-0.011 to +0.133) | Post-selection sensitivity |

**Note:** Positive ΔAUC values favor the FAQ-based or FAQ-added model. The Locked FAQ Trio rows evaluate the screen defined as any difficulty in at least 1 of the 3 activities comprising finances/checkbook, shopping and games/hobbies, without reopening screen selection. Primary selected screen analyses estimate the held-out performance of the prespecified FAQ screen selection procedure, not the apparent performance of a rule chosen using the full dataset. The Locked FAQ Trio rows provide internal post selection sensitivity analyses and do not constitute external validation.
**Abbreviations:** AUC, area under the receiver operating characteristic curve; BH, Benjamini-Hochberg; CI, confidence interval; FAQ, Functional Activities Questionnaire; MMSE, Mini-Mental State Examination; PET, positron emission tomography; SUVR, standardized uptake value ratio.

### Fixed Locked FAQ Trio Sensitivity Analyses

Once the Locked FAQ Trio selected, a similar nested repeated cross-validation internal analysis of the fixed Locked FAQ Trio only, was evaluated and confirmed downstream interpretability. In Block A, the predictably of 3-year progression to dementia in Base + Locked FAQ Trio showed a higher discrimination with AUC 0.823 compared with 0.776 for Base + entorhinal tau PET SUVR (ΔAUC, 0.047; 95% CI, –0.061 to 0.147). In Block B, the Locked FAQ Trio showed an AUC of 0.822 compared with 0.636 for Base + MMSE (ΔAUC, 0.186; 95% CI, 0.071 to 0.311). Similarly, in incremental prognostic value analyses, the Locked FAQ Trio showed directionally positive incremental discrimination beyond entorhinal tau PET SUVR (ΔAUC, 0.079; 95% CI, 0.005 to 0.156) and beyond entorhinal tau PET SUVR and MMSE score (ΔAUC, 0.059; 95% CI, –0.011 to 0.133). Fixed Locked FAQ Trio rows are shown above in Table 2 and early progressor sensitivity appears in eTable 5.

### Clinical Risk Separation and Operating Characteristics

The Locked FAQ Trio effectively captured escalations per converter confirming its use case as a front-end triage of prognosis prediction risk to dementia in MCI patients. In the entorhinal tau PET SUVR predictor analysis Block A, the Locked FAQ Trio was prevalent in 37.6% (59/157) of participants and captured 84.4% (27/32) of progressors, or 45.8% (27/59) of participants with any difficulty in at least 1 of the 3 Locked FAQ Trio activities (or screen-positive) progressed to Alzheimer disease dementia versus 5.1% (5/98) of screen-negative participants, yielding a 84.4% sensitivity, 74.4% specificity, 45.8% PPV and 94.9% NPV. Similar results were observed in the MMSE scores analysis Block B, where the Locked FAQ Trio was prevalent in 37.3% (57/153) participants and captured 83.9% (26/31) of progressors, or 45.6% (26/57) of screen-positive participants progressed to dementia versus 5.2% (5/96) of screen-negative participants, yielding a 83.9% sensitivity, 74.6% specificity, 45.6% PPV and 94.8% NPV. Descriptive Cox full follow-up survival models were adjusted for age at tau PET visit, sex recorded in ADNI, education and APOE ε4. These full follow-up survival studies achieved comparable results, with 35.3% (41/116) and 35.1% (40/114) of screen-positive progressing to dementia versus 6.5% (13/199) and 6.5% (12/184) of the screen-negative participants with adjusted hazard ratios 7.46 and 8.19 in the entorhinal tau PET SUVR cohort and in the MMSE scores cohort, respectively. In both entorhinal tau PET SUVR and MMSE scores cohorts, the Kaplan-Meier curves showed early and sustained separation, suggesting that a Locked FAQ Trio front-end triage clearly captures both near-term and longer-term risks of progression. Operating characteristics for the fixed Locked FAQ Trio rules are shown in Table 3.

**Table 3.** Operating Characteristics and Clinical Risk Separation of Fixed Locked FAQ Trio in Primary Benchmarking Analysis Aim 1.

| Block | Rule | Screen+ | Prog if + | Prog if - | Sensitivity | Specificity | PPV / NPV |
| --- | --- | --- | --- | --- | --- | --- | --- |
| A | Locked FAQ Trio | 59/157 (37.6%) | 27/59 (45.8%) | 5/98 (5.1%) | 84.4% (67.2-94.7%) | 74.4% (65.8-81.8%) | 45.8% (32.7-59.2%) / 94.9% (88.5-98.3%) |
| A | Companion FAQ Trio | 85/157 (54.1%) | 31/85 (36.5%) | 1/72 (1.4%) | 96.9% (83.8-99.9%) | 56.8% (47.6-65.6%) | 36.5% (26.3-47.6%) / 98.6% (92.5-100.0%) |
| A | FAQ total $\geq 6$ | 36/157 (22.9%) | 20/36 (55.6%) | 12/121 (9.9%) | 62.5% (43.7-78.9%) | 87.2% (80.0-92.5%) | 55.6% (38.1-72.1%) / 90.1% (83.3-94.8%) |
| B | Locked FAQ Trio | 57/153 (37.3%) | 26/57 (45.6%) | 5/96 (5.2%) | 83.9% (66.3-94.5%) | 74.6% (65.9-82.0%) | 45.6% (32.4-59.3%) / 94.8% (88.3-98.3%) |
| B | Companion FAQ Trio | 82/153 (53.6%) | 30/82 (36.6%) | 1/71 (1.4%) | 96.8% (83.3-99.9%) | 57.4% (48.1-66.3%) | 36.6% (26.2-48.0%) / 98.6% (92.4-100.0%) |
| B | FAQ total $\geq 6$ | 35/153 (22.9%) | 19/35 (54.3%) | 12/118 (10.2%) | 61.3% (42.2-78.2%) | 86.9% (79.6-92.3%) | 54.3% (36.6-71.2%) / 89.8% (82.9-94.6%) |

**Note:** Operating characteristics are descriptive and calculated on common model-specific denominators. The Locked FAQ Trio is defined as any difficulty in at least 1 of 3 activities comprising finances/checkbook, shopping and games/hobbies. The Companion FAQ Trio is defined as any difficulty in at least 1 of 3 activities comprising forms/papers, shopping and remembering appointments/medications/holidays. FAQ total score $\geq 6$ is a summed-score severity rule.
**Abbreviations:** FAQ, Functional Activities Questionnaire; GE1, FAQ item score of Greater or Equal to 1; GE2, FAQ item score of Greater or Equal to 2; NPV, negative predictive value; PPV, positive predictive value.

We also evaluated a fixed companion GE1 screen, defined as any difficulty in at least 1 of the 3 activities of forms/papers, shopping and remembering appointments/medications/holidays (Companion FAQ Trio) for sensitivity. While it was not the top-performing screen in the nested cross-validation selection model, it remains clinically informative and captured nearly all progressors or 96.9% (31/32) in the entorhinal tau PET cohort. In Block A, screen-positive prevalence was 54.1% (85/157) with 36.5% (31/85) of those screen-positive participants progressing within 3 years versus 1.4% (1/72) in screen-negative participants and an adjusted hazard ratio of 5.54. The Companion FAQ Trio showed similar results in Block B and clear separation in Cox full follow-up analysis as well as in Kaplan-Meier curves. The Companion FAQ Trio captured more progressors than the Locked FAQ Trio but would also send more patients to downstream triage. This supports the use of the Locked FAQ Trio, more selective, for front end triage while the Companion FAQ Trio offers a higher-sensitivity but lower-specificity triage option with 45.8% and 36.5% of screen-positive progressing respectively. Full follow-up Kaplan-Meier risk separation is shown in eFigure 2 and survival discrimination is summarized in eTable 6.

**Figure 2.**
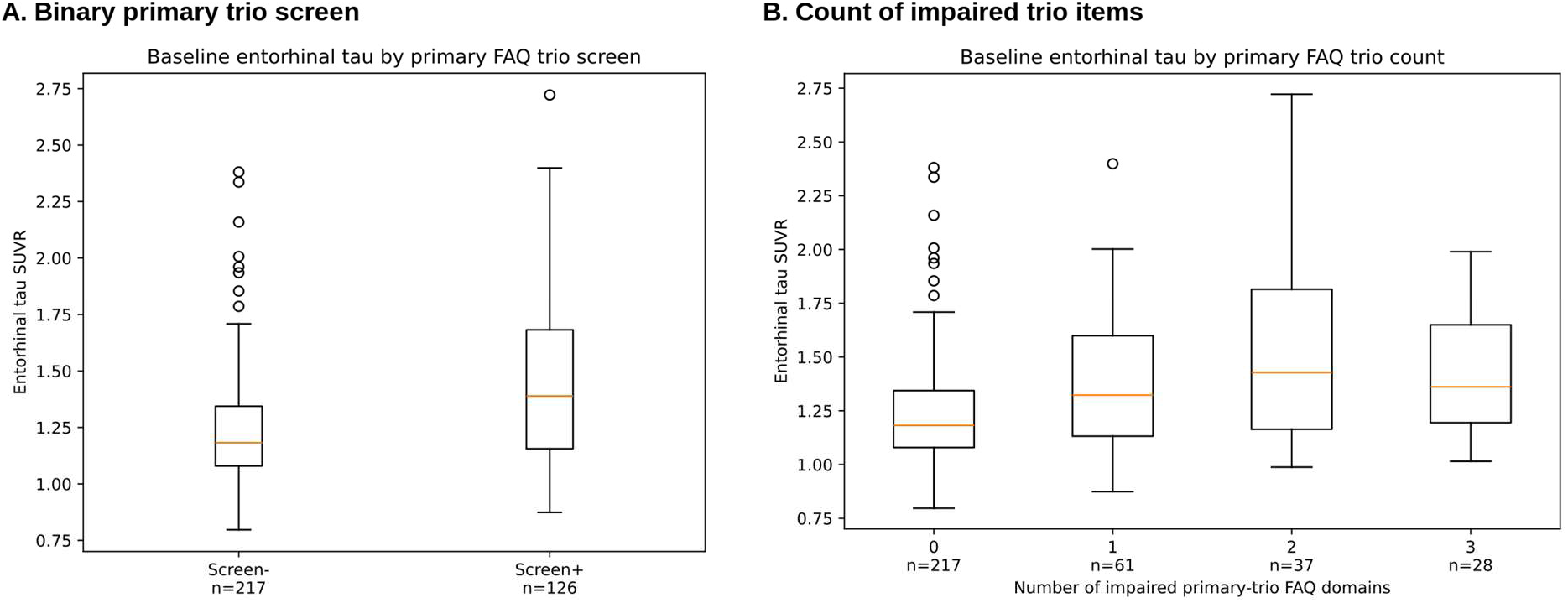
Baseline Entorhinal Tau PET SUVR by Locked FAQ Trio Status and Item Count. **Note:** The Locked FAQ Trio was defined as any difficulty in at least 1 of 3 activities comprising finances/checkbook, shopping and games/hobbies. The binary panel compares entorhinal tau PET SUVR by Locked FAQ Trio status. The count panel shows entorhinal tau PET SUVR across 0, 1, 2 and 3 positive Locked FAQ Trio activities. The count panel supports descriptive interpretation only, the observed pattern suggests enrichment after the first 1 to 2 positive activities, with possible plateauing rather than a strictly monotonic dose-response across all 3 activities. The small all-3-positive subgroup limits inference about incremental biological information from the 3^rd^ activity. **Abbreviations:** FAQ, Functional Activities Questionnaire; PET, positron emission tomography; SUVR, standardized uptake value ratio.

### Survival, Calibration, Decision Curves, Timing and Robustness

Censoring-aware survival analyses were robust across all sensitivity analyses. Survival discrimination analyses were consistent with the primary binary results and reinforced the tau PET cohort findings. In cross-validated follow-up survival discrimination analyses, the Locked FAQ Trio signal increased Harrell C index from 0.694 to 0.775 in Block A and from 0.637 to 0.779 in Block B with ΔC estimates of +0.081 and +0.142, respectively. In survival horizon analyses, the Locked FAQ Trio remained numerically higher to tau PET (Block A) at 2 years and at 3 years, which are the most relevant clinically, while there were attenuations at five years.

Predicted and observed 3-year progression rates were aligned for most participants, especially in the range where most predicted probabilities fell, with calibration plots broadly acceptable showing no notable miscalibration across the central range of predicted risk. Uncertainty was wider at the extremes, the observed rates were noisier given fewer participants and observations in those ranges. While there are no universal thresholds to refer to, the exploratory decision curve analyses were supportive, showing a higher net benefit for the Locked FAQ Trio across several plausible escalation or referral triage thresholds, notably from 0.10 to 0.30. Supporting survival discrimination, decision curve and calibration displays are provided in eTable 6 and eFigures 2, 3 and 4.

The primary GE1 signal was generally preserved over sensitivity analyses, including re-anchoring the baseline at the latest available predictor date (IndexMax), restricting to predictors obtained within a 30-day window (Window30), removing of APOE ε4 from the adjustment set, coding missing FAQ items as unimpaired and simplifying under the prespecified parsimony tie rule. Exclusions of early progressors were analyzed at 90 and 180 days, they suggested that the FAQ signal was strongest nearer to the clinical progression from MCI to dementia boundary. Early progressor and timing sensitivity analyses are summarized in eTable 5 and eTable 12 and timing distributions are shown in eFigure 9.

### Primary Aim 2: Locked FAQ Trio Versus UPENN Plasma p-tau217

In the primary benchmarking analysis Aim 2, we conducted a similar prognosis prediction study comparing the same Locked FAQ Trio to p-tau217 and tau PET predictors. The study derived from a smaller UPENN p-tau217 subset of 66 participants with MCI at baseline, endpoint-known 3-year progression status as well as predictors and covariate data available. In this cohort, 19.7% (13/66) of participants had progressed to dementia within 3 years, consistent with the cohort used in Aim 1. Given the limited size of the plasma cohort, we did not reopen the FAQ screen discovery and carried the Locked FAQ Trio from Aim 1 screen selection.

In the comparison of standalone predictors for 3-year progression prediction risk, using a repeated cross-validation base-covariates model, p-tau217 emerged as the highest discrimination among fixed models. In descending order of AUC in standalone base-covariates models: Base + p-tau217 (AUC, 0.890; Brier, 0.081), Base + entorhinal tau PET SUVR (AUC, 0.798; Brier, 0.139), Base + Locked FAQ Trio (AUC, 0.749; Brier, 0.131) and Base (AUC, 0.601; Brier, 0.151). Adding predictors combinations to the model, Base + p-tau217 remains the best predictor of progression, while adding Locked FAQ Trio or entorhinal tau PET SUVR to p-tau217 appeared as close runner ups with Base + p-tau217 + Locked FAQ Trio AUC of 0.881 and Base + p-tau217 + entorhinal tau PET SUVR AUC of 0.878 AUC. Fixed model plasma results are shown in Table 4, Panel A.

**Table 4.** – Panel A. Primary Benchmarking Analysis Aim 2: Fixed Model Discrimination in the UPENN Plasma Model-Specific Subset.

| Model | n (events) | AUC | Brier |
| --- | --- | --- | --- |
| Base + p-tau217 | 66 (13) | 0.890 | 0.081 |
| Base + Locked FAQ Trio + p-tau217 | 66 (13) | 0.881 | 0.086 |
| Base + tau PET + p-tau217 | 66 (13) | 0.878 | 0.090 |
| Base + tau PET + Locked FAQ Trio + p-tau217 | 66 (13) | 0.862 | 0.092 |
| Base + tau PET | 66 (13) | 0.798 | 0.130 |
| Base + Locked FAQ trio | 66 (13) | 0.749 | 0.131 |
| Base | 66 (13) | 0.604 | 0.151 |

**Note:** All fixed models use the same 66 participants denominator including 13 progressors. P-tau217 values are sourced from the UPENN plasma context used for the primary plasma benchmarking analysis.
**Abbreviations:** AUC, area under the receiver operating characteristic curve; FAQ, Functional Activities Questionnaire; PET, positron emission tomography; p-tau217, phosphorylated tau 217; UPENN, University of Pennsylvania.

**Table 4.**
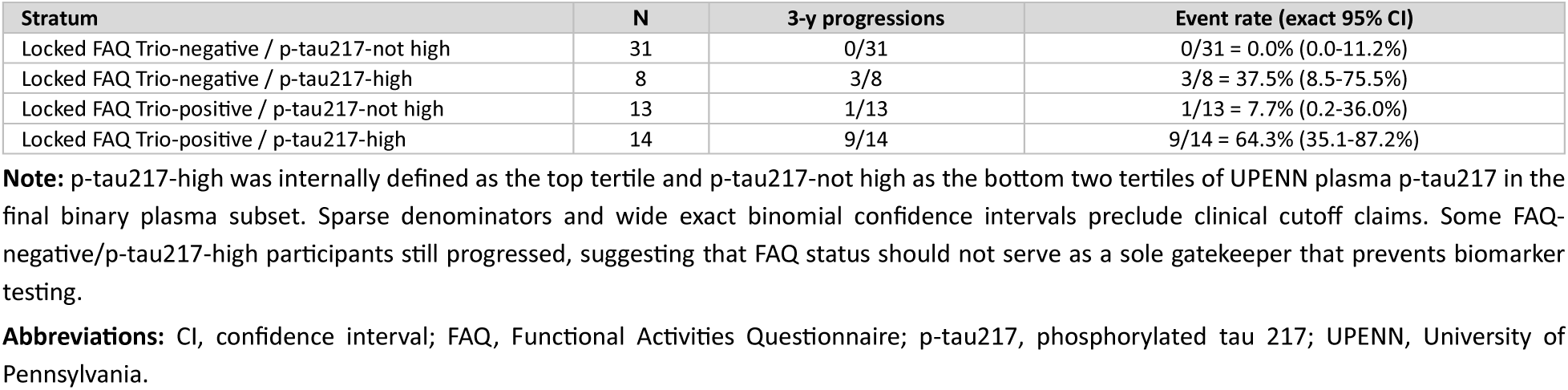
– Panel B. FAQ-by-p-tau217 Risk Strata in the UPENN Plasma Model-Specific Subset.

| Stratum | N | 3-y progressions | Event rate (exact 95% CI) |
| --- | --- | --- | --- |
| Locked FAQ Trio-negative / p-tau217-not high | 31 | 0/31 | 0/31 = 0.0% (0.0-11.2%) |
| Locked FAQ Trio-negative / p-tau217-high | 8 | 3/8 | 3/8 = 37.5% (8.5-75.5%) |
| Locked FAQ Trio-positive / p-tau217-not high | 13 | 1/13 | 1/13 = 7.7% (0.2-36.0%) |
| Locked FAQ Trio-positive / p-tau217-high | 14 | 9/14 | 9/14 = 64.3% (35.1-87.2%) |

**Note:** p-tau217-high was internally defined as the top tertile and p-tau217-not high as the bottom two tertiles of UPENN plasma p-tau217 in the final binary plasma subset. Sparse denominators and wide exact binomial confidence intervals preclude clinical cutoff claims. Some FAQ-negative/p-tau217-high participants still progressed, suggesting that FAQ status should not serve as a sole gatekeeper that prevents biomarker testing.
**Abbreviations:** CI, confidence interval; FAQ, Functional Activities Questionnaire; p-tau217, phosphorylated tau 217; UPENN, University of Pennsylvania.

A combination of the Locked FAQ Trio with p-tau217 strata, internally defined as p-tau-high for the top tertile and p-tau217-not high for the two bottom tertiles, offered clinically relevant risk separations. Notably, none (0 of 31) of the MCI participants in the lower risk cohort progressed, defined as Locked FAQ Trio screen-negative/p-tau217-not high. In addition, 64.3% (9 of 14) of MCI participants in the higher risk cohort progressed, defined as Locked FAQ Trio-positive/p-tau217-high. There were 37.5% (3 of 8) participants in the Locked FAQ Trio-negative/p-tau 217-high subset who still progressed, showing that while FAQ can contextualized p-tau217, it should not be used as a substitute for biological biomarkers and patients should not be excluded from downstream testing when clinical concerns remain. This reinforced p-tau217 second position in our suggested 3-step triage hierarchy. FAQ-by-p-tau217 risk strata are shown in Table 4, Panel B and in Figure 1.

In exploratory same denominator analyses combining plasma biomarkers, adding plasma Aβ42/40 to UPENN p-tau217 (AUC, 0.888) or a quartet including p-tau217, Aβ42/40, GFAP, and NfL (AUC, 0.866), did not materially improve the 3-year progression prediction risk, showing the restraint around multiprotein modeling. These results are exploratory and should not be generalized to other plasma platforms, regulatory assays or clinical cutoffs. Secondary and exploratory plasma marker models are summarized in eTable 8.

### Biological Context: Entorhinal Tau PET and Amyloid PET

The Locked FAQ Trio was also biologically aligned with Alzheimer disease biology. The Locked FAQ Trio screen-positive participants had substantially higher baseline entorhinal tau PET SUVR than those screen-negative (mean SUVR 1.443 versus 1.241; Mann-Whitney U test, P = 5.37 x 10-8), this association was preserved after adjusting for age at tau PET anchor visit, sex, education, APOE ε4, MMSE, amyloid status and Centiloid. Count-based analyses also showed enrichment with mean entorhinal tau PET SUVR increasing from 1.241 in participants with none of the 3 items within the Locked FAQ Trio, to 1.401 with 1 item impaired and 1.522 with 2 items impaired. In all-3 activities impaired it remained elevated at 1.428 but the small subgroup size limits inferences on whether the 3^rd^ item adds incremental biology context, but support a brief screen selection rather than a benefit to extend the screen across many additional activities. The tau-burden pattern is shown in Figure 2 and summarized in eTable 7.

Amyloid PET sensitivity analyses informed the biological context rather than a direct progression risk predictor comparison. Positivity for the Locked FAQ Trio was more common in Aβ+ than Aβ– participants (46.1% versus 29.7%), but the screen remained informative after Aβ adjustment and within the Aβ+ subgroup. Among Aβ+ participants with endpoint-known 3-year progression status, progression to Alzheimer disease dementia occurred in 55.6% (20/36) of screen-positive participants versus 16.2% (6/37) of those screen-negative. Aβ-adjusted logistic odds ratios remained large and Aβ-adjusted Cox hazard ratios ranged from 6.72 to 8.25 across the main model families. The formal tests for screen-by-amyloid interaction were not conclusive, supporting the interpretation that the Locked FAQ Trio aligns with, but is not reducible to, Aβ status alone. Amyloid and tau biological context are summarized in eTable 7, with biomarker context displays in eFigures 7 and eFigure 8.

### p-tau181/p-tau231 Source Availability Audit

Conducting an audit of the p-tau181/p-tau231 sources, we identified a FNIH plasma pTau file containing PTAU_181 and PTAU_231 as well as an alternative UGOT p-tau181 file containing PLASMAPTAU181. The exact FNIH PTAU_181 source matched 21 participants anchored at tau within the prespecified 90-day window. The strict p-tau217 and broader endpoint-known source-availability checks remained too sparse for modeling and contained no 3-year progressors in the exact FNIH PTAU_181 common denominators. For these reasons, we did not report p-tau181 prognostic models or risk stratification analyses. This audit did not evaluate the Roche Elecsys pTau181 assay, its clinical cutoff or its intended clinical pathway. The p-tau181/p-tau231 source availability audit is summarized in eTable 9.

### Exploratory Diagnostic Resource-Use Scenarios

We translated the observed operating characteristics into a per 1,000 MCI individuals context to analyze diagnostic resource-use scenarios. We considered that the per-1000 values would scale testing volume only and the observed progressor count would remain tied to the source denominators.

Locked FAQ Trio versus entorhinal tau PET SUVR scan (Aim 1): considering the Block A of 157 MCI participants (including 32 progressors), a Locked FAQ Trio-first pathway would send approximately 376 patients to tau PET scan per 1,000 MCI patients, avoiding 624 tau PET scan visits compared to a universal tau PET-first pathway, and yet capturing 27 of the 32 observed progressors. The Companion FAQ Trio (forms/papers, shopping and remembering appointments/medications/holidays) would send approximately 541 patients to tau PET scan, avoiding 459 tau PET scan visits and capturing 31 of the 32 progressors to dementia. The FAQ total sum score of ≥6 threshold would send approximately 229 patients to tau PET scan, avoiding 771 scans and capturing 20 of the 32 progressors.

Locked FAQ Trio versus plasma p-tau217 (Aim 2): in the 66 MCI participants (including 13 progressors) in the plasma subset, we evaluated several internal nonclinical scenarios. In the top-tertile scenario in which p-tau217-high status would trigger a tau PET scan escalation, approximatively 333 patients per 1,000 would be sent downstream to tau PET scan, capturing 12 of the 13 progressors. In a higher sensitivity scenario defined as Locked FAQ Trio-positive or p-tau217-high, approximately 530 participants would be sent to tau PET scan, yet capturing all progressors. In a higher specificity scenario requiring both Locked FAQ Trio-positive and p-tau217-high, approximately 212 participants would be sent downstream and capture 9 of the 13 observed progressors. These scenarios quantify diagnostic testing volume and illustrative tradeoffs only, they do not constitute cost-effectiveness, cost-benefit or treatment-access analyses. Diagnostic resource use scenarios are summarized in eTable 10.

## Discussion

### Principal Findings

In this prognosis prediction risk study, an informant-reported functional signal supports near-term prognostic triage in MCI. A leakage-controlled cross-validation selection model identified the Locked FAQ Trio, a 3-item FAQ-derived stable screen defined as any difficulty of at least 1 of the 3 activities comprising finances/checkbook, shopping and games/hobbies. In held-out base-covariates analyses, the Locked FAQ Trio showed numerically higher discrimination than entorhinal tau PET SUVR and statistically significantly higher discrimination than MMSE score.

Plasma p-tau217 served as the strongest scalable biological predictor of progression risk in this study. UPENN plasma p-tau217 showed the highest discrimination among fixed models tested in the plasma subset.

The Locked FAQ Trio value lies in clinical triage, it provides a scalable low burden tool that may help clinicians prioritize and contextualize the near-term progression risk among MCI patients. The Locked FAQ Trio should not be interpreted as a substitute for biological biomarker nor as a gatekeeper preventing further biomarker testing.

Together, these findings support a staged prognostic triage pathway starting with a brief informant-based functional assessment informing the front-end clinical risk context, then plasma p-tau217 providing a scalable biological refinement before amyloid confirmatory PET or CSF testing, tau PET staging, specialist counseling, repeat blood testing or longitudinal reassessment can follow according to the clinical question.

### Clinical-Biological Interpretation

There is an inherent circularity we appreciate given that functional decline captured in a brief FAQ itself contributes to the dementia diagnosis. However, there is evidence to suggest that this Locked FAQ Trio is sensitive to changes beyond inexorable proximity to a dementia diagnosis. First, this trio aligns with both Entorhinal tau uptake and Aβ positivity separate from 3-year progression status. Entorhinal tau SUVR signal changes with item count (1.241, 1.401, and 1.522 across 0, 1 or 2 items, plateauing at 3 items, as one would expect with a short screen instead of a dose-response pattern). These relationships remain independent of any clinical transition. Second, hazard ratios stayed quite large (6.72–8.25) when adjusting for Aβ, indicating that any present signal is likely not reflective of Aβ alone, leaving it a valid measure independent of traditional biological associations. Together, these findings support this measure as one that may reflect an underlying biological vulnerability across the path to dementia rather than a reflection of the clinical diagnostic criteria. This reflects the principle of staged biomarker democratization, where we can assemble evidence of mixed and broad signals, each providing insight, to some extent, into likely underlying biological processes and not simply an echoing of a broader, more thorough clinical assessment. Such an argument allows us to interpret the change in early progressor AUC (+0.005) as expected behavior in a measure best suited for triaging earlier rather than later progression to dementia. This is an an ideal use case for a near zero-cost, low-burden tool that can be deployed at the precipice of imminent conversion.

### Blood-Biomarker Implementation and Non-interchangeable Regulatory Use Cases

The plasma findings are aligned with a growing literature that positions p-tau217 as the leading scalable blood-based biomarker [7], [8], [9], [10], [11], [13], [14], [15]. All plasma analyses in this study were based on same-source plasma biomarker specification in a small ADNI denominator and should not be generalized to all ratio measures, commercial algorithms, clinical cutoffs or regulatory assays. The internal p-tau217 strata analyses used a top tertile threshold, this is not a clinical threshold and should not be treated as such, those analyses support exploratory risk stratification only. Recent work indicates that p-tau217 values and cutoffs may vary with comorbidities such as kidney function, body mass index and anemia [42]. While this does not weaken p-tau217 analyses, it highlights why biomarker pathways should incorporate clinical context, assay calibration and downstream confirmatory strategies.

Recent regulatory developments reinforce the assay-specific interpretation and the non-interchangeability of blood-based biomarkers. In 2025, the FDA cleared Lumipulse G pTau217/β-Amyloid 1-42 Plasma Ratio to aid identification of amyloid pathology in symptomatic adults evaluated in specialized care settings [12]. Later in 2025, the FDA also cleared Elecsys Phospho-Tau (181P) Plasma assay, but for a different use case than the p-tau217/amyloid-ratio pathway. The plasma p-tau181 assay is intended to aid initial assessment of Alzheimer disease and other causes of cognitive decline in symptomatic adults aged 55 years or older and the FDA decision summary states that its performance has not been established for predicting progression to dementia or monitoring response to therapy [43]. The plasma analyses in this study should not be interpreted as evaluation of neither Lumipulse nor Elecsys assays, their prespecified cutoffs or their intended use pathways.

### Tau PET Interpretation

This study benchmarked FAQ-derived screens against the biological imaging comparator entorhinal tau PET SUVR measure obtained in ADNI and did not test other regions of interest of tau PET scans. Entorhinal tau PET SUVR is a powerful staging and prognostic tool with recent literature supporting its strong prognostic value in MCI, especially when the clinical question requires biological staging, regional tau burden, trial enrichment or separation of biological disease stage from clinical syndrome [16], [17]. The FAQ-derived study evaluated a different question: whether an informant-reported functional change can help identify MCI patients more likely to approach a clinically meaningful transition to Alzheimer disease dementia. The appropriate interpretation is that a brief informant-reported functional signal showed discrimination in the same observed range of entorhinal tau PET SUVR for near-term clinical progression.

### Implications For Treatment Pathways

The FDA relatively recently approved anti-amyloid disease-modifying therapies addressing underlying pathology, and the accessibility of those therapies are increasing the need for staged diagnostic triage workflow, yet they did not change the endpoint of the primary analysis in this study, which is progression from MCI to Alzheimer disease dementia. The current labels of two such therapies, lecanemab and donanemab, require amyloid confirmation before treatment and the indication is limited to patients with MCI or mild dementia [18], [19], [20], [21]. This distinction is clinically important because those treatments uptake bottleneck is not only whether Alzheimer disease biology can be detected, but also whether early symptomatic patients can be identified, staged, confirmed and referred before functional decline advances beyond the treatment indication window. While a FAQ-derived screen cannot establish treatment eligibility, it may help inform clinicians decision making as to who needs more urgent biomarker evaluation, specialist counseling or closer follow-ups.

### Diagnostic Stewardship

The staged diagnostic triage suggested in this study also has stewardship benefits given the capacity constraints of PET imaging, CSF testing, plasma biomarkers, specialist counseling and MRI safety monitoring. The low burden front-end additional layer in the diagnosis triage framework does not require any assay or imaging and thus may expand the proportion of MCI patients entering the prognosis prediction diagnosis pathway. This additional front-end layer of triage does not devalue advanced biomarker innovation, but rather may help deploy such innovation more efficiently.

### Exploratory Diagnostic Resource-Use Scenarios

The exploratory resource-use scenarios illustrate a practical triage performance translation into volume implications per 1,000 MCI patients and should not be interpreted as a formal economic model.

In the entorhinal tau PET SUVR primary analysis (Aim 1), a FAQ-derived first pathway could reduce low-yield tau PET scan volume in the ADNI samples, but they would also miss some progressors depending on the FAQ-derived screen selected sensitivity-specificity tradeoff. A universal tau PET scan pathway would send all 1,000 MCI patients to tau PET scan, while a Locked FAQ Trio-first pathway would send approximately 376 of 1,000 MCI patients downstream to tau PET, thus avoiding approximately 624 PET scans and yet capturing 27 of the 32 progressors within 3 years. A Companion FAQ Trio-first would prioritize 541 of 1,000 MCI patients to obtain a tau PET scan, “avoiding” approximately 459 scans and capturing 31 of the 32 progressors. In another scenario, a FAQ total score ≥6-first would send prioritize 229 of 1,000 MCI patients to obtain a tau PET scan, avoiding approximately 771 scans and capturing 20 of the 32 progressors.

In the smaller plasma p-tau217 subset primary analysis (Aim 2), combined Locked FAQ Trio and p-tau217 scenarios illustrate how biological biomarkers and functional signals could complement one another. A universal p-tau217 testing with tau PET scan escalation in the p-tau217-high MCI population would prioritize approximately 333 of 1,000 MCI patients to obtain a downstream tau PET scan, yet capturing 12 of the 13 progressors within 3 years. A higher-sensitivity strategy using either Locked FAQ Trio-positive or p-tau217-high status would prioritize approximately 530 of 1,000 MCI patients to obtain a tau PET scan while capturing all 13 progressors. A higher-specificity scenario requiring both Locked FAQ Trio-positive and p-tau217-high status would prioritize approximately 212 of 1,000 MCI patients to obtain a tau PET scan but captured only 9 of the 13 progressors. We observed that 3 of the 8 MCI participants that are Locked FAQ Trio-negative and p-tau217-high progressed, this emphasizes that FAQ-derived screens should not be used as a stand-alone gatekeeper to prevent further biomarker testing.

These illustrative exploratory diagnostic analyses quantified testing volume but did not model treatment effects, drug acquisition costs, infusion costs, MRI monitoring, amyloid-related imaging abnormality (ARIA) management, utilities, patient out-of-pocket costs or workflow integrations. These analyses do not establish that a staged diagnostic triage workflow improves cost-effectiveness or treatment benefit, but rather that this suggested triage pathway may help reduce low-yield imaging volume and help prioritize biomarker escalation.

### Strengths and Limitations

This study presents several strengths: it used a clearly defined clinical progression assessment, separated screen discovery from independent performance evaluation through nested cross-validation, evaluated repeated out-of-fold predictions at the participant level, benchmarked FAQ-derived screens against a biological entorhinal tau PET SUVR imaging comparator, retained p-tau217 as the primary plasma comparator, separated Aim 1 and Aim 2 denominators. It also evaluated sensitivity analyses around subgroups, calibration, decision-curve, survival, timing, early-progressor exclusions and endpoint-ascertainment IPW. It also explicitly treated amyloid PET, tau PET and resource-use as additional biological context rather than primary comparators.

Several limitations need to be emphasized: the ADNI cohort is highly characterized but does not fully represent primary care, community, rural, under-resourced, low-education and culturally diverse memory-evaluations populations [30], [31]. More than half of the tau PET scan visit anchored cohort lacked ascertainable progression status endpoint within 3 years for the primary binary analyses. The study used internal cross-validation rather than external validation. The plasma p-tau217 subset comprised only 66 participants, including 13 progressors within 3 years. The internal p-tau217-high threshold was internally defined as upper tertile but does not represent a clinical cutoff. The entorhinal tau PET SUVR comparator should not be generalized to all tau PET methods. Given that functional decline contributes to clinical dementia diagnosis, the Locked FAQ Trio signal partly captures proximity to clinical transition. p-tau181 and p-tau231 could not be evaluated in this study given the sparse overlap among source-availability audit, the tau PET scan visit anchor, the endpoint-known 3-year progression status and UPENN p-tau217 data availability. The resource-use scenario remained exploratory, U.S.-centric and scenario-based.

### Future Directions

Future studies should externally validate the Locked FAQ Trio in more diverse memory-clinic, primary care and community populations. They should also recalibrate the plasma biomarker combinations by assay and settings and evaluate differential item functioning across education, culture, sex, digital literacy and disability strata. Further studies should also test whether repeated FAQ-derived assessments improve prediction beyond a single baseline measure and test electronic health record integration or web-based pre-visit triage workflows. Implementation studies should measure not only discrimination but also referral yield, time to biomarker confirmation, false reassurance, diagnostic cascade burden, patient and care-partner experience, specialist capacity, equity of access and workflow feasibility.

### Conclusions

In this ADNI secondary prognostic prediction study of MCI participants, a leakage-controlled FAQ-derived screen selection procedure repeatedly identified a stable stand-alone 3-item signal defined as any reported difficulty in at least 1 of 3 activities comprising finances/checkbook, shopping and games/hobbies, with item-level convergence on the same functional cluster. The Locked FAQ Trio supported near-term clinical risk stratification that can be obtained before PET, CSF, or plasma testing and may help organize biomarker workflows. It is aligned with entorhinal tau burden, amyloid burden and plasma p-tau217 biological contexts. In the plasma subset, p-tau217 showed the highest cross-validated discrimination among the fixed models tested, and FAQ-by-p-tau217 strata further separated progression risk prediction, stressing plasma p-tau217 role as a central biological comparator that can provide scalable biological refinement. This staged diagnostic triage approach is compatible with the direction of the field as biological Alzheimer disease diagnosis is becoming more precise, blood-based biomarkers are becoming more scalable, and tau PET scans remain powerful for staging and prognosis. Clinical function adds a relevant layer as patients and clinicians need to know not only whether Alzheimer disease pathology is present, but also how close the patient may be to a clinically meaningful transition.

These findings support a staged prognostic-triage framework where low burden informant-reported functional information can contextualize and prioritize biomarker evaluation. But it should not replace plasma biomarkers, amyloid confirmation, CSF testing, tau PET staging or treatment-eligibility assessments. FAQ should not be interpreted as a sole gatekeeper preventing further biomarker evaluations, especially when clinical concern remains, and external validation in a larger and more representative cohort is necessary. However, the core finding remains: a rigorously examined, statistically thorough analysis converges on the utility of a three-question informant-reported assessment that can be answered in minutes, with discrimination on par with advanced neuroimaging biomarkers. This demonstrates how careful analysis of existing data, in light of biological relevance, can fulfill a promise of biomarker democratization, where it is not a substitute for the current state of necessary assessments, but as a carefully distilled path navigable for broad populations of high-risk individuals (and their caregivers) who would be otherwise unable or reluctant to enter it.

## Article Information

### CRediT authorship contribution statement

Juliette Lafille: Conceptualization (equal), Methodology (lead), Software (lead), Validation (lead), Formal analysis (lead), Investigation (lead), Data curation (lead), Visualization (lead), Writing: original draft (lead), Writing: review & editing (equal). Frank Provenzano: Conceptualization (equal), Methodology (supporting), Validation (supporting), Supervision (lead), Writing: review & editing (equal).

### Data Sharing Statement

Source data used in this study were obtained from the Alzheimer’s Disease Neuroimaging Initiative (ADNI) database and are available to qualified investigators through ADNI data-access and data-use procedures. Derived analytic code and run-stamped output tables may be made available by the corresponding author upon reasonable request.

### Reporting Guideline

We used TRIPOD+AI as the primary prediction-model reporting framework, STROBE for observational-cohort context, PROBAST/PROBAST+AI for self-audit, and STARD-style operating characteristics for descriptive interpretation only.

## Data Availability

https://adni.loni.usc.edu/data-samples/adni-data/

## Abbreviations

Aβ: amyloid-β
ADNI: Alzheimer’s Disease Neuroimaging Initiative
APOE ε4: apolipoprotein E ε4
ARIA: amyloid-related imaging abnormality
AUC: area under the receiver operating characteristic curve
BH: Benjamini-Hochberg
CSF: cerebrospinal fluid
Companion FAQ Trio: any informant-reported impairment difficulty in at least 1 of the 3 activities comprising forms/papers, shopping and remembering appointments/medications/holidays
FAQ: Functional Activities Questionnaire
GE1: FAQ item score Greater or Equal to 1, ≥1
GE2: FAQ item score Greater or Equal to 2, ≥2
GFAP: glial fibrillary acidic protein
IPW: inverse-probability weighting
Locked FAQ Trio: any informant-reported impairment difficulty in at least 1 of the 3 activities comprising finances/checkbook, shopping and games/hobbies
MCI: mild cognitive impairment
MMSE: Mini-Mental State Examination
MRI: magnetic resonance imaging
NfL: neurofilament light chain
NPV: negative predictive value
PET: positron emission tomography
PPV: positive predictive value
p-tau: phosphorylated tau
SUVR: standardized uptake value ratio

## Supplemental Tables

**eTable 1.** Cohort Flow, Missingness and Timing Alignment.

| Component | Value | Comment |
| --- | --- | --- |
| Known 3-y status | 163/350 (46.6%) | Primary denominator loss |
| FAQ games/hobbies | 342/350 usable | 2.3% missing |
| FAQ finances/checkbook | 344/350 usable | 1.7% missing |
| FAQ shopping | 344/350 usable | 1.7% missing |
| FAQ total | 341/350 usable | 2.6% missing |
| APOE ε4 | 325/350 usable | 7.1% missing |
| MMSE | 332/350 usable | 5.1% missing |
| Entorhinal tau PET | 347/350 usable | 0.9% missing |
| Tau PET to FAQ interval | Median 15.0 d (IQR 5.2 to 44.0) | Within 90 d: 324/350 (92.6%) |
| MMSE relative to tau PET | Median -47.5 d (IQR -82.0 to -27.8) | Within 90 d: 266/332 (80.1%) |

**Note:** Component-specific denominators are shown where available. Endpoint missingness, not predictor missingness, drove most binary analyses denominator loss.

**eTable 2.** Functional Activities Questionnaire Items, Response Options and Harmonized Scoring.

**Panel A. FAQ item domains used in the screen library**
| Analysis variable | Domain label | Activity assessed |
| --- | --- | --- |
| FAQFINAN | Finances/checkbook | Writing checks, paying bills, or balancing a checkbook |
| FAQFORM | Forms/papers | Assembling tax records, business affairs, or other papers |
| FAQSHOP | Shopping | Shopping alone for clothes, household necessities, or groceries |
| FAQGAME | Games/hobbies | Playing a game of skill or working on a hobby |
| FAQBEVG | Heating water/stove | Heating water, making coffee, or turning off the stove |
| FAQMEAL | Meal preparation | Preparing a balanced meal |
| FAQEVENT | Current events | Keeping track of current events |
| FAQTV | TV/reading comprehension | Paying attention to and understanding television, a book, or a magazine |
| FAQREM | Remembering | Remembering appointments, family occasions, holidays, or medications |
| FAQTRAVL | Travel/transportation | Traveling outside the neighborhood, driving, or arranging transportation |

**Panel B. Worksheet response options and harmonized scoring**
| Response option | Worksheet item score | ADNI raw code | Harmonized analysis score |
| --- | --- | --- | --- |
| Normal | 0 | 0 | 0 |
| Never did, but could do now | 0 | 1 | 0 |
| Never did, would have difficulty now | 1 | 2 | 1 |
| Has difficulty, but does by self | 1 | 3 | 1 |
| Requires assistance | 2 | 4 | 2 |
| Dependent | 3 | 5 | 3 |

**Note:** FAQ item responses were harmonized to the 0-to-3 item score scale before analysis. GE1 was defined as Greater or Equal harmonized item score $\geq 1$ , indicating any reported difficulty. GE2 was defined as Greater or Equal harmonized item score $\geq 2$ , indicating at least moderate difficulty. FAQ total score was calculated as the strict sum of the 10 harmonized item scores when all item values were available, with possible range 0 to 30. ADNI datasets may store item responses as 0-to-5 categorical response codes corresponding to the 6 response options shown.
**Abbreviations:** ADNI, Alzheimer's Disease Neuroimaging Initiative; FAQ, Functional Activities Questionnaire; GE1, FAQ item score Equal or Greater to 1; GE2, FAQ item score Equal or Greater to 2.

**eTable 3.** Screen Selection Stability and GE2 Sensitivity.

| Threshold | Block | Most frequent screen | Share |
| --- | --- | --- | --- |
| GE1 | A. FAQ vs tau PET | Finances/checkbook + shopping + games/hobbies | 31% |
| GE1 | B. FAQ vs MMSE | Finances/checkbook + shopping + games/hobbies | 32% |
| GE1 | C. Add FAQ to tau PET | Forms/papers + shopping + games/hobbies + meal | 18% |
| GE1 | D. Add FAQ to tau PET + MMSE | Forms/papers + shopping + remembering | 22% |
| GE2 | A. FAQ vs tau PET | FAQ total score $\geq 6$ | 98% |
| GE2 | B. FAQ vs MMSE | FAQ total score $\geq 6$ | 100% |
| GE2 | C. Add FAQ to tau PET | FAQ total score $\geq 6$ | 72% |
| GE2 | D. Add FAQ to tau PET + MMSE | FAQ total score $\geq 6$ | 75% |

**Note:** Shares indicate the proportion of outer-loop resamples selecting the listed rule. GE2 concentrated on FAQ total score $\geq 6$ rather than on an ultra-brief item combination.

**eTable 4.**
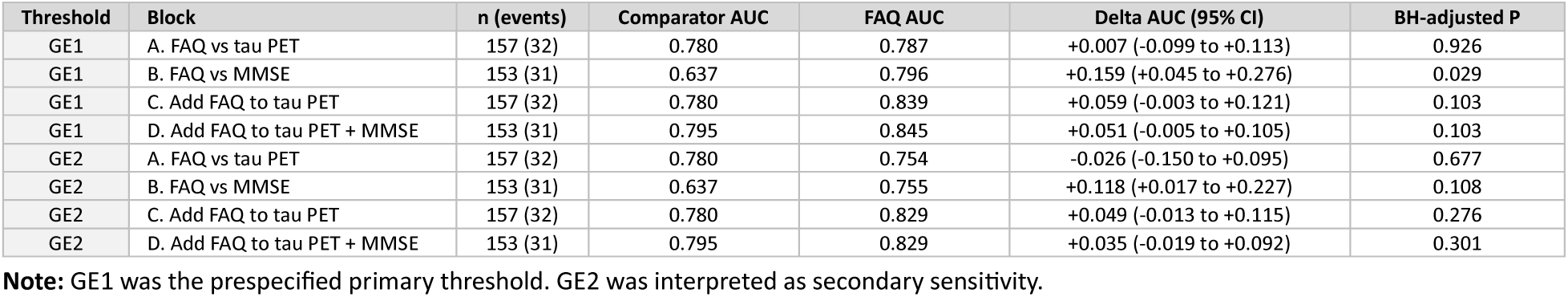
Full Primary Benchmarking Analysis Aim 1 Discrimination for GE1 and GE2.

**eTable 5.** Fixed Locked FAQ Trio Early Progressor and IPW Sensitivity.

| Analysis | Block | n (events) | Comparator AUC | FAQ AUC | Delta AUC (95% CI) |
| --- | --- | --- | --- | --- | --- |
| Exclude ≤ 90 d progressors | A. Locked FAQ Trio vs entorhinal tau PET SUVR | 156 (31) | 0.799 | 0.826 | +0.027 (-0.074 to +0.122) |
| Exclude ≤ 90 d progressors | B. Locked FAQ Trio vs MMSE score | 153 (31) | 0.627 | 0.821 | +0.194 (+0.083 to +0.313) |
| Exclude ≤ 90 d progressors | C. Add Locked FAQ Trio to entorhinal tau PET SUVR | 156 (31) | 0.799 | 0.860 | +0.061 (-0.001 to +0.131) |
| Exclude ≤ 90 d progressors | D. Add Locked FAQ Trio to entorhinal tau PET SUVR + MMSE score | 153 (31) | 0.790 | 0.855 | +0.065 (-0.008 to +0.135) |
| Exclude ≤ 180 d progressors | A. Locked FAQ Trio vs entorhinal tau PET SUVR | 155 (30) | 0.816 | 0.821 | +0.005 (-0.088 to +0.093) |
| Exclude ≤ 180 d progressors | B. Locked FAQ Trio vs MMSE score | 152 (30) | 0.639 | 0.815 | +0.176 (+0.058 to +0.302) |
| Exclude ≤ 180 d progressors | C. Add Locked FAQ Trio to entorhinal tau PET SUVR | 155 (30) | 0.816 | 0.863 | +0.047 (-0.011 to +0.108) |
| Exclude ≤ 180 d progressors | D. Add Locked FAQ Trio to entorhinal tau PET SUVR + MMSE score | 152 (30) | 0.810 | 0.857 | +0.046 (-0.017 to +0.110) |
| IPW diagnostics | Endpoint-known model | known 153;<br>unknown 144 |  |  | Median weight 1.85;<br>95th pct 2.60; ESS 147.8 |

**Note:** Early progressor rows evaluate the sensitivity to exclusion of very early transitions. IPW rows are supportive endpoint ascertainment sensitivity analyses.
**Abbreviations:** IPW, inverse-probability weighting; PET, positron emission tomography.

**eTable 6.**
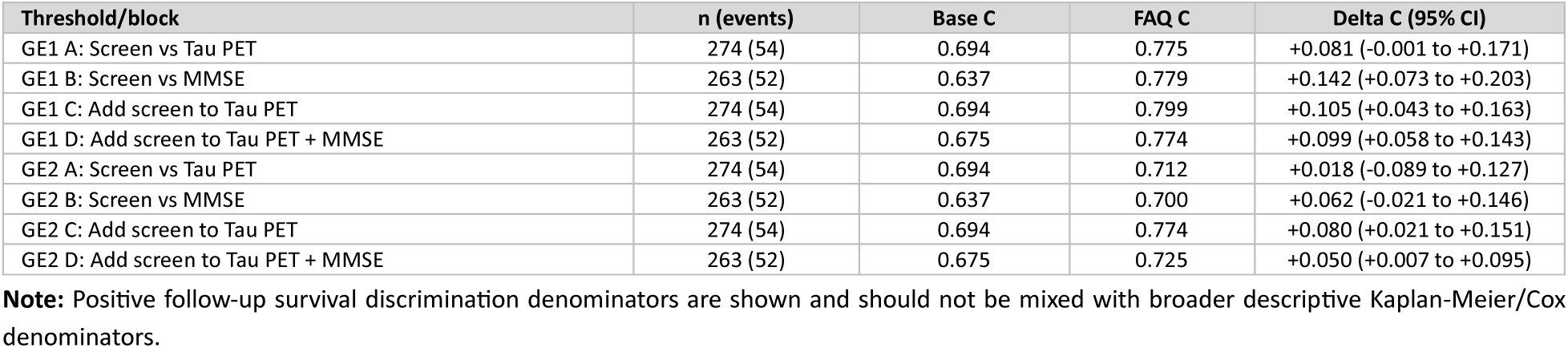
Survival Discrimination.

**eTable 7.** Tau and Amyloid PET Biological Context.

| Component | Result |
| --- | --- |
| Amyloid PET matching | 303/350 matched; median gap 9.0 days |
| Screen positivity if amyloid-negative | 44/148 (29.7%) |
| Screen positivity if amyloid-positive | 70/152 (46.1%) |
| Amyloid-positive progression if screen-negative | 6/37 (16.2%) |
| Amyloid-positive progression if screen-positive | 20/36 (55.6%) |
| Amyloid-adjusted OR range | 8.99-13.15 |
| Amyloid-adjusted HR range | 5.64-8.25 |
| Entorhinal Tau PET SUVR by trio count 0 | n=217; mean 1.241 (0.253) |
| Entorhinal Tau PET SUVR by trio count 1 | n=61; mean 1.401 (0.314) |
| Entorhinal Tau PET SUVR by trio count 2 | n=37; mean 1.522 (0.423) |
| Entorhinal Tau PET SUVR by trio count 3 | n=28; mean 1.428 (0.310) |

**Note:** Amyloid PET context uses component-specific denominators. Entorhinal tau PET SUVR summaries are biological-context analyses, not diagnostic biomarker replacement analyses.
**Abbreviations:** PET, positron emission tomography; SUVR, standardized uptake value ratio.

**eTable 8.** Secondary and Exploratory Plasma Marker Models.

| Model | n (events) | AUC | Brier |
| --- | --- | --- | --- |
| Base | 66 (13) | 0.605 | 0.152 |
| Locked FAQ Trio | 66 (13) | 0.743 | 0.131 |
| p-tau217 | 66 (13) | 0.893 | 0.080 |
| p-tau217 + Aβ42/40 | 66 (13) | 0.888 | 0.081 |
| 4-marker composite | 66 (13) | 0.866 | 0.108 |
| Locked FAQ Trio + p-tau217 | 66 (13) | 0.880 | 0.084 |
| Locked FAQ Trio + p-tau217 + Aβ42/40 | 66 (13) | 0.878 | 0.087 |
| Locked FAQ Trio + 4-marker composite | 66 (13) | 0.866 | 0.106 |
| Paired plasma Aβ42/40 add-on: p-tau217 + Aβ42/40 minus p-tau217 |  |  | -0.001 (-0.012 to +0.007) |
| Paired plasma Aβ42/40 add-on: Locked FAQ Trio + p-tau217 + Aβ42/40 minus Locked FAQ Trio + p-tau217 |  |  | -0.001 (-0.015 to +0.011) |

**Note:** These marker specification rows are secondary/exploratory and should not replace the primary p-tau217 fixed model comparison. Paired plasma Aβ42/40 is evaluated only as an add-on to UPENN plasma p-tau217 on the same model-specific denominator.
**Abbreviations:** AUC, area under the receiver operating characteristic curve; FAQ, Functional Activities Questionnaire; p-tau217, phosphorylated tau 217; UPENN, University of Pennsylvania.

**eTable 9.**
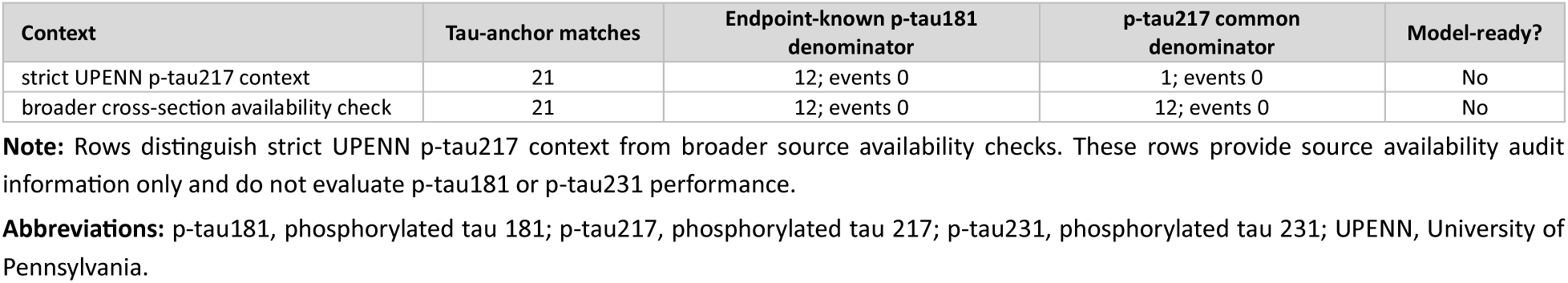
P-tau181/p-tau231 Source Availability Audit.

**eTable 10.** Exploratory Diagnostic Resource-Use Scenarios per 1,000 MCI Individuals.

| Context | Scenario | Plasma /1000 | Tau PET /1000 | Avoided PET /1000 | Captured / outcome | Sensitivity |
| --- | --- | --- | --- | --- | --- | --- |
| Aim 1 | Universal tau PET benchmark | 0 | 1000 | 0 | 32/32 | 100.0% |
| Aim 1 | Locked FAQ Trio-first | 0 | 376 | 624 | 27/32 | 84.4% |
| Aim 1 | Companion FAQ Trio-first | 0 | 541 | 459 | 31/32 | 96.9% |
| Aim 1 | FAQ total score ≥6 | 0 | 229 | 771 | 20/32 | 62.5% |
| Aim 2 | Universal tau PET benchmark within plasma subset | 0 | 1000 | 0 | 13/13 | 100.0% |
| Aim 2 | Universal p-tau217, tau PET if p-tau217 high | 1000 | 333 | 667 | 12/13 | 92.3% |
| Aim 2 | Universal p-tau217 plus Locked FAQ Trio; tau PET if Locked FAQ Trio-positive or p-tau217-high | 1000 | 530 | 470 | 13/13 | 100.0% |
| Aim 2 | Sequential Locked FAQ Trio-first, p-tau217 only if Locked FAQ Trio-positive, tau PET if both positive | 409 | 212 | 788 | 9/13 | 69.2% |
| Aim 2 | Universal p-tau217 plus Locked FAQ Trio, tau PET if both Locked FAQ Trio-positive and p-tau217 high | 1000 | 212 | 788 | 9/13 | 69.2% |
| Aim 2 | Locked FAQ Trio-negative / p-tau217-nohigh descriptive stratum | 1000 |  |  | 0/31 |  |

**Note:** Rows are derived from source-specific observed operating characteristics and scaled to 1,000 MCI individuals within each context. Scenarios rows are not cost-effectiveness, cost-benefit nor treatment-access analyses. FAQ-only scenarios use Primary Benchmarking Analysis Aim 1 denominators and plasma-derived scenarios use Primary Benchmarking Analysis Aim 2 denominators.
**Abbreviations:** FAQ, Functional Activities Questionnaire; MCI, mild cognitive impairment; PET, positron emission tomography; p-tau217, phosphorylated tau 217.

**eTable 11.** Descriptive Subgroup and Role Sensitivity Profiles.

| Denominator | Subgroup | Level | n (events) | Screen+ | Sensitivity | Specificity | NPV |
| --- | --- | --- | --- | --- | --- | --- | --- |
| Block A | Sex recorded in ADNI | Women | 56 (15) | 37.5% | 86.7% | 80.5% | 94.3% |
| Block A | Sex recorded in ADNI | Men | 101 (17) | 37.6% | 82.4% | 71.4% | 95.2% |
| Block A | Age | <75 y | 85 (16) | 35.3% | 87.5% | 76.8% | 96.4% |
| Block A | Age | ≥75 y | 72 (16) | 40.3% | 81.2% | 71.4% | 93.0% |
| Block A | Education | <16 y | 48 (9) | 50.0% | 88.9% | 59.0% | 95.8% |
| Block A | Education | ≥16 y | 109 (23) | 32.1% | 82.6% | 81.4% | 94.6% |
| Block A | APOE ε4 | Noncarrier | 92 (18) | 35.9% | 83.3% | 75.7% | 94.9% |
| Block A | APOE ε4 | Carrier | 65 (14) | 40.0% | 85.7% | 72.5% | 94.9% |
| Block B | Sex recorded in ADNI | Women | 55 (15) | 38.2% | 86.7% | 80.0% | 94.1% |
| Block B | Sex recorded in ADNI | Men | 98 (16) | 36.7% | 81.2% | 72.0% | 95.2% |
| Block B | Age | <75 y | 83 (15) | 33.7% | 86.7% | 77.9% | 96.4% |
| Block B | Age | ≥75 y | 70 (16) | 41.4% | 81.2% | 70.4% | 92.7% |
| Block B | Education | <16 y | 47 (9) | 48.9% | 88.9% | 60.5% | 95.8% |
| Block B | Education | ≥16 y | 106 (22) | 32.1% | 81.8% | 81.0% | 94.4% |
| Block B | APOE ε4 | Noncarrier | 91 (18) | 36.3% | 83.3% | 75.3% | 94.8% |
| Block B | APOE ε4 | Carrier | 62 (13) | 38.7% | 84.6% | 73.5% | 94.7% |

**Note:** Subgroup rows are descriptive and not powered for formal subgroup interaction or differential item functioning inference.
**Abbreviations:** APOE ε4, apolipoprotein E ε4; FAQ, Functional Activities Questionnaire; NPV, negative predictive value; PPV, positive predictive value.

**eTable 12.**
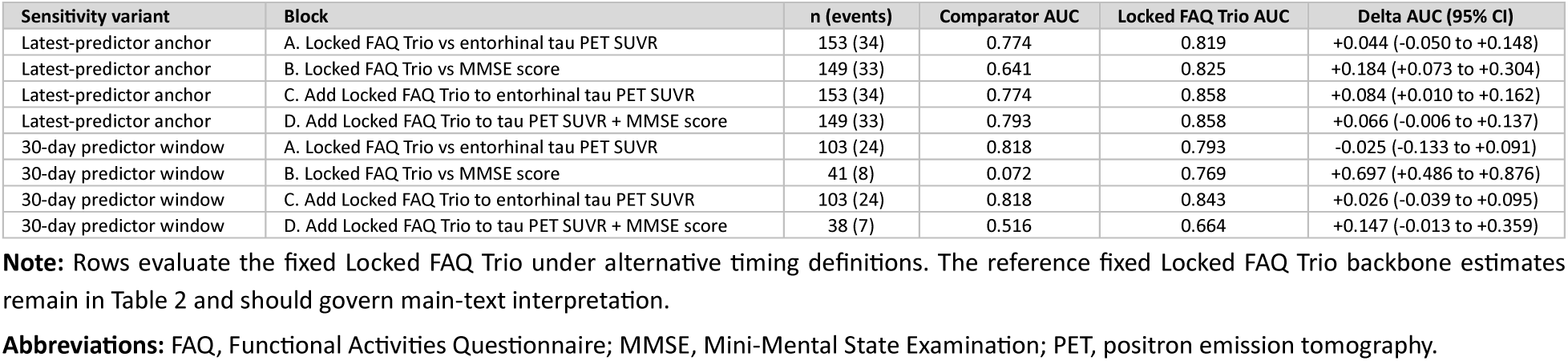
Supplemental Fixed Locked FAQ Trio Timing Sensitivity Analyses.

| Sensitivity variant | Block | n (events) | Comparator AUC | Locked FAQ Trio AUC | Delta AUC (95% CI) |
| --- | --- | --- | --- | --- | --- |
| Latest-predictor anchor | A. Locked FAQ Trio vs entorhinal tau PET SUVR | 153 (34) | 0.774 | 0.819 | +0.044 (-0.050 to +0.148) |
| Latest-predictor anchor | B. Locked FAQ Trio vs MMSE score | 149 (33) | 0.641 | 0.825 | +0.184 (+0.073 to +0.304) |
| Latest-predictor anchor | C. Add Locked FAQ Trio to entorhinal tau PET SUVR | 153 (34) | 0.774 | 0.858 | +0.084 (+0.010 to +0.162) |
| Latest-predictor anchor | D. Add Locked FAQ Trio to tau PET SUVR + MMSE score | 149 (33) | 0.793 | 0.858 | +0.066 (-0.006 to +0.137) |
| 30-day predictor window | A. Locked FAQ Trio vs entorhinal tau PET SUVR | 103 (24) | 0.818 | 0.793 | -0.025 (-0.133 to +0.091) |
| 30-day predictor window | B. Locked FAQ Trio vs MMSE score | 41 (8) | 0.072 | 0.769 | +0.697 (+0.486 to +0.876) |
| 30-day predictor window | C. Add Locked FAQ Trio to entorhinal tau PET SUVR | 103 (24) | 0.818 | 0.843 | +0.026 (-0.039 to +0.095) |
| 30-day predictor window | D. Add Locked FAQ Trio to tau PET SUVR + MMSE score | 38 (7) | 0.516 | 0.664 | +0.147 (-0.013 to +0.359) |

**Note:** Rows evaluate the fixed Locked FAQ Trio under alternative timing definitions. The reference fixed Locked FAQ Trio backbone estimates remain in Table 2 and should govern main-text interpretation.
**Abbreviations:** FAQ, Functional Activities Questionnaire; MMSE, Mini-Mental State Examination; PET, positron emission tomography.

## Supplemental Figures

**eFigure 1.**
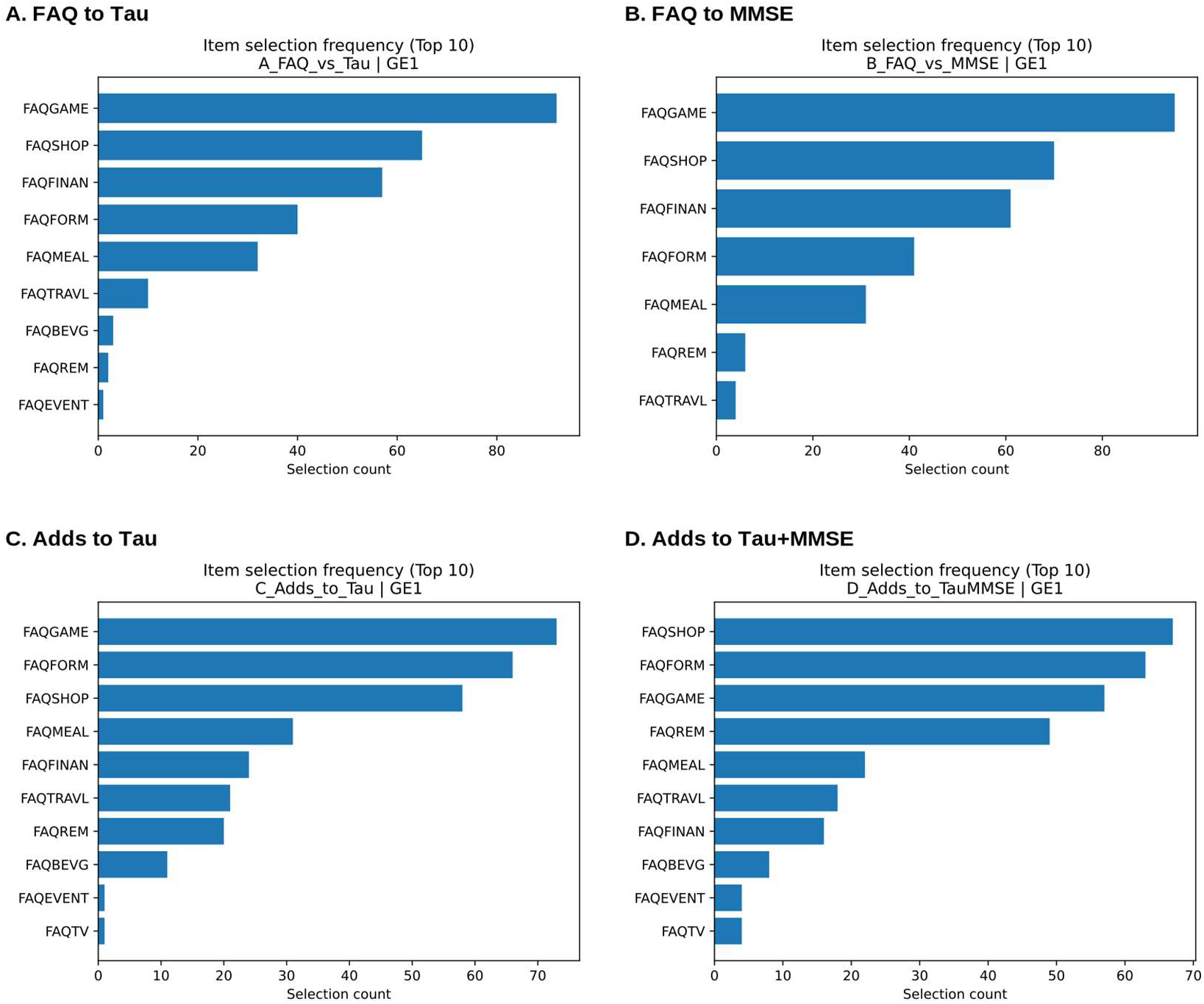
GE1 Item Selection Frequencies Across Repeated Nested Cross-Validation. **Note:** Bars show how often each FAQ item was included in selected GE1 screens across 100 outer-loop resamples per block. GE1 was defined as Greater or Equal harmonized FAQ item score ≥1, indicating any reported difficulty. The display shows the concentration of selection around a small functional cluster rather than diffuse selection across all FAQ items. **Abbreviations:** FAQ, Functional Activities Questionnaire; GE1, FAQ item score of Equal or Greater to ≥1.

**eFigure 2.**
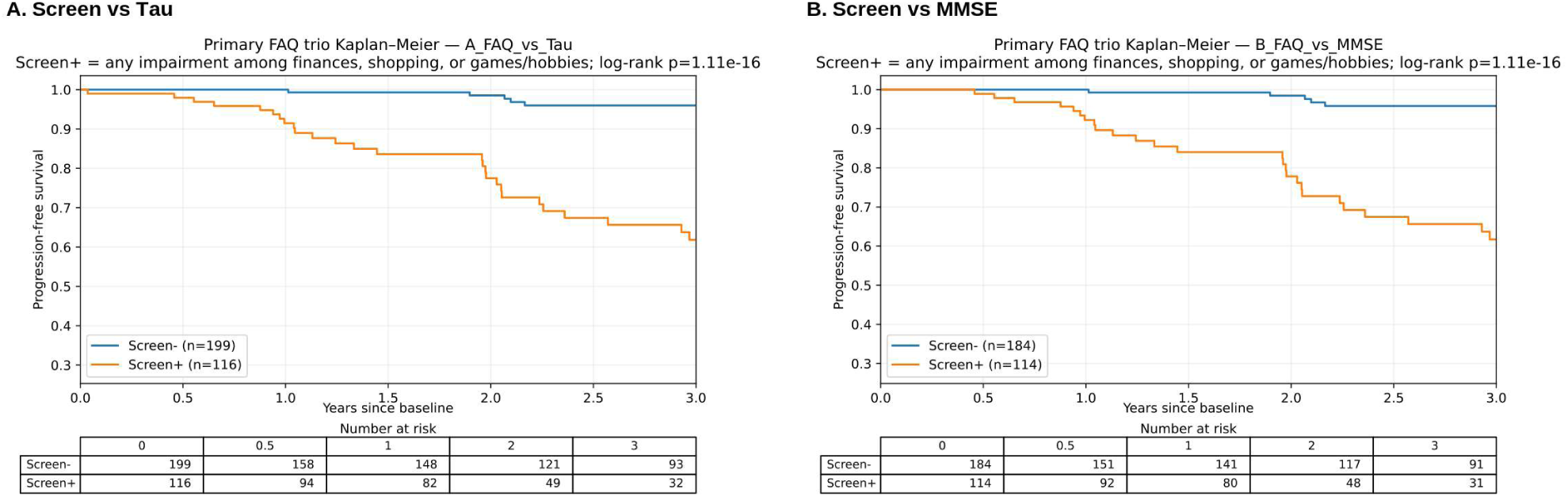
Kaplan-Meier Curves for the Locked FAQ Trio. **Note:** Kaplan-Meier curves show full follow-up progression-free probability by Locked FAQ Trio status in the entorhinal tau PET SUVR and MMSE score available cohorts. Screen-positive status indicates reported difficulty in at least 1 of 3 activities comprising finances/checkbook, shopping and games/hobbies. **Abbreviations:** FAQ, Functional Activities Questionnaire; KM, Kaplan-Meier; MMSE, Mini-Mental State Examination; PET, positron emission tomography.

**eFigure 3.**
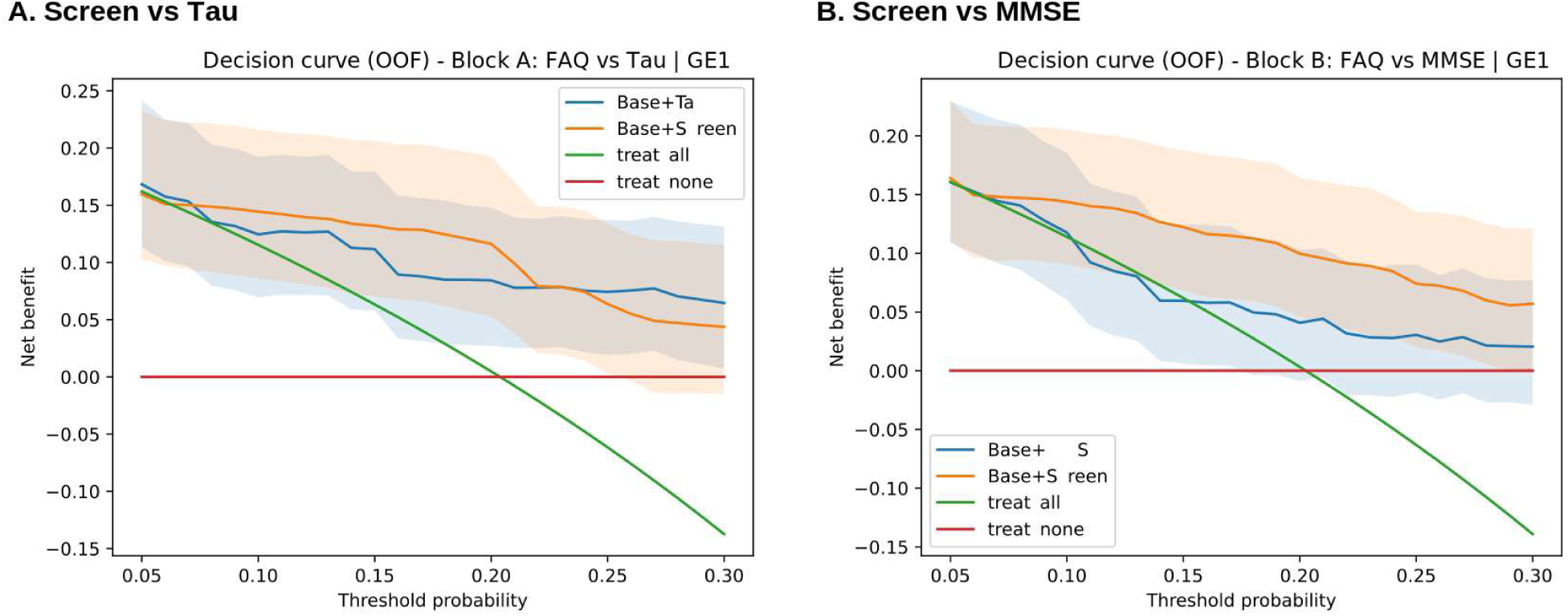
Decision Curve Analysis for GE1 Models in Blocks A and B. **Note:** Decision curves plot net benefit across threshold probabilities for the primary GE1 comparison models. These analyses are exploratory and should not define a universal biomarker-escalation or referral threshold. **Abbreviations:** FAQ, Functional Activities Questionnaire; GE1, FAQ item score of Greater or Equal to ≥1; MMSE, Mini-Mental State Examination.

**eFigure 4.**
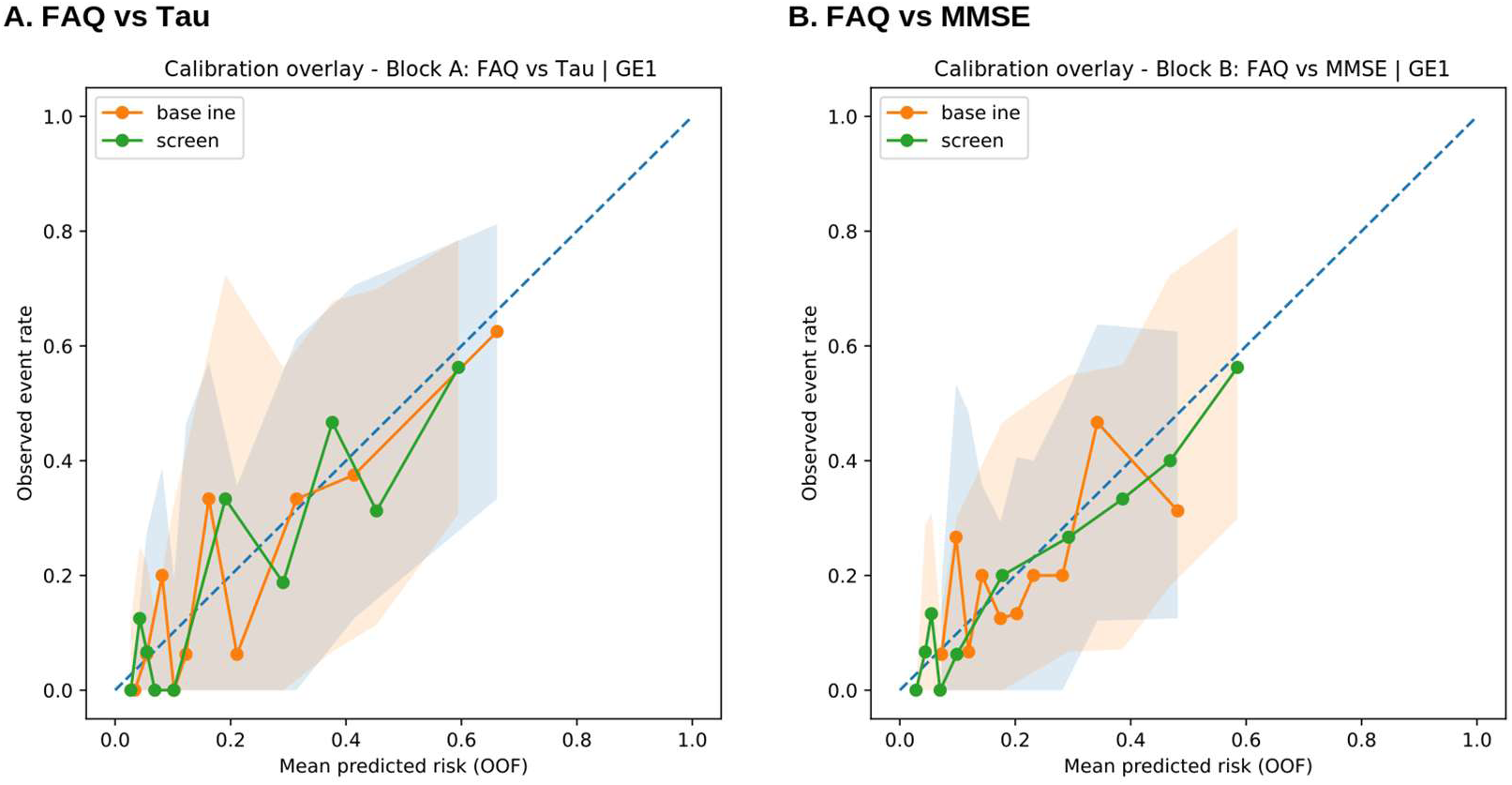
Calibration Overlays for Primary GE1 Comparison Models. **Note:** Calibration plots compare participant-level out-of-fold predicted risk with observed 3-year progression risk. The plots support visual assessment of calibration across the central predicted risk range and show wider uncertainty at the extremes. **Abbreviations:** FAQ, Functional Activities Questionnaire; GE1, FAQ item score Equal or Greater to ≥1.

**eFigure 5.**
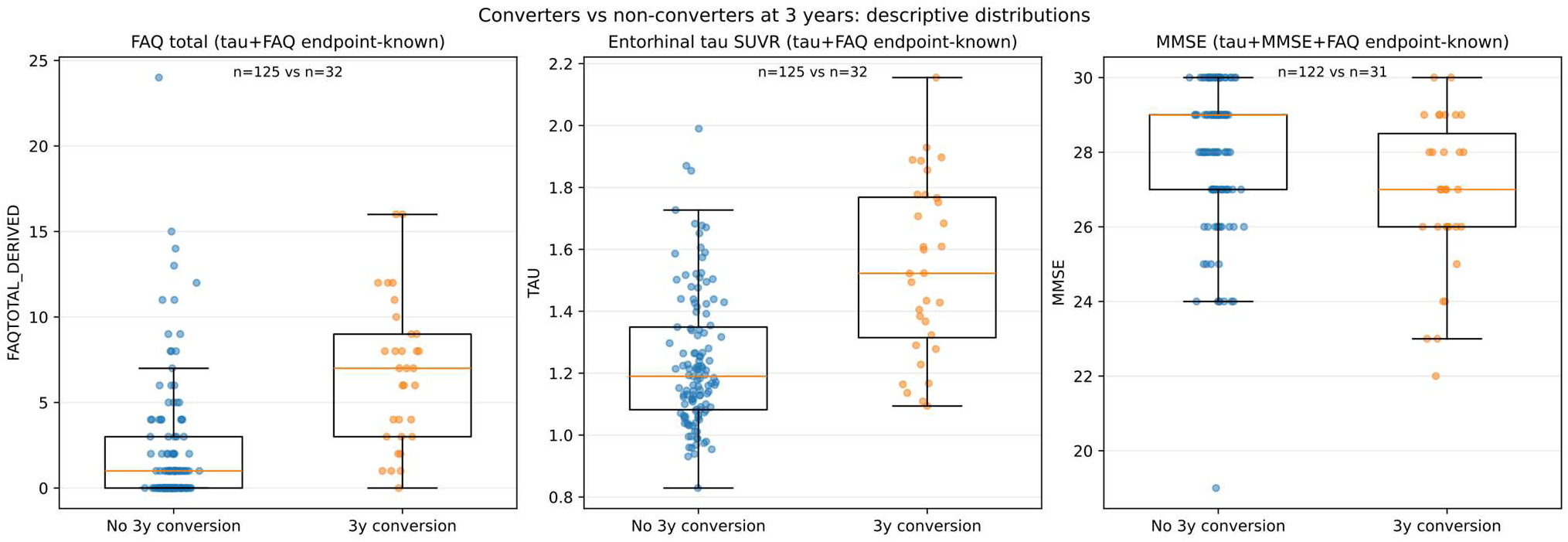
Descriptive Baseline Differences Between 3-Year Progressors and Non-Progressors. **Note:** Panels summarize baseline clinical and biomarker contrasts between participants who did and did not progress within 3 years among those with known-endpoint status. These displays provide descriptive context for the prediction analyses. **Abbreviations:** FAQ, Functional Activities Questionnaire; MMSE, Mini-Mental State Examination; PET, positron emission tomography.

**eFigure 6.**
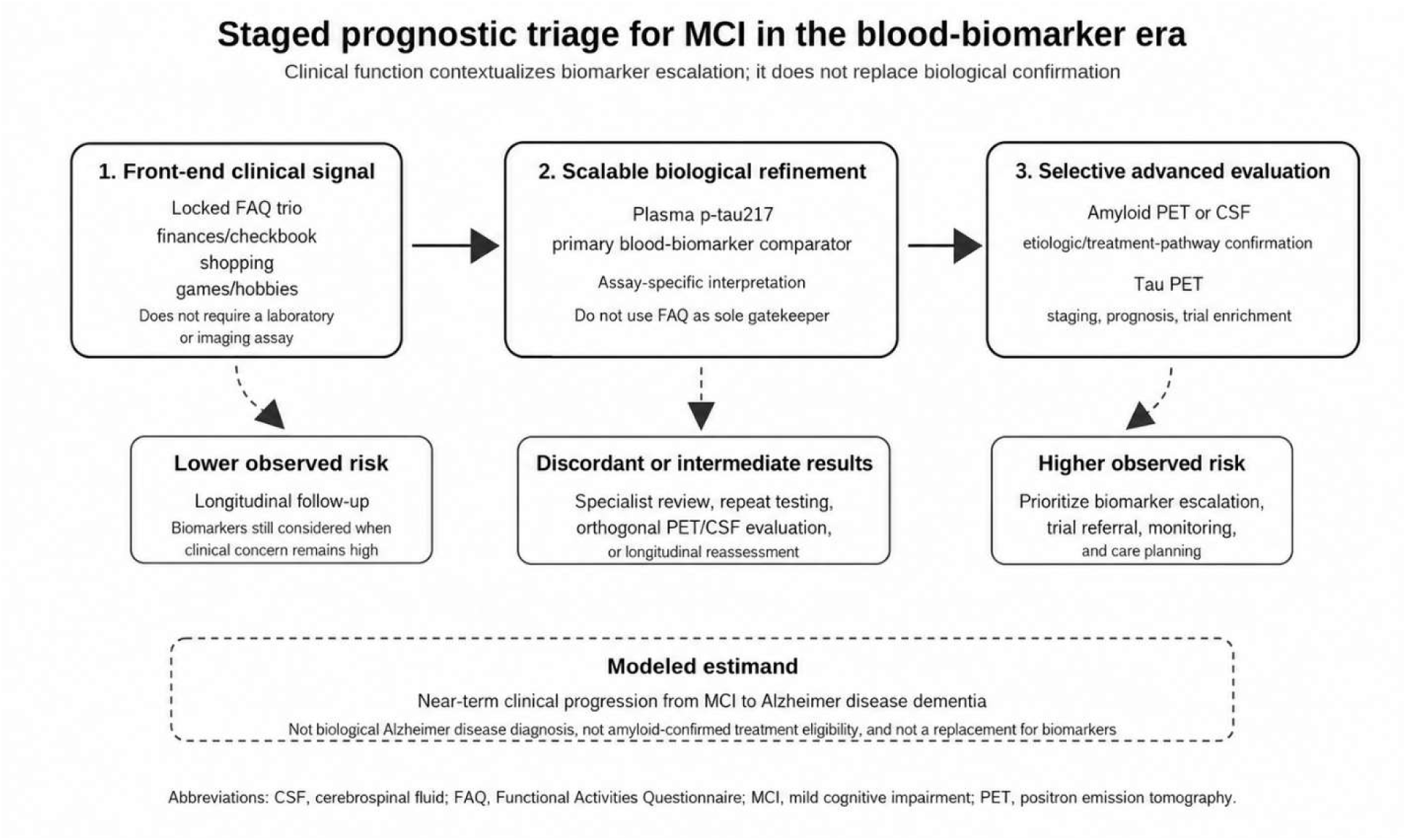
Conceptual Staged Prognostic Triage Workflow. **Note:** The schematic illustrates how brief informant-reported function, plasma p-tau217 and selective PET or CSF evaluation may answer complementary clinical and biological questions. This is a conceptual workflow figure, not a validated clinical algorithm. FAQ-derived screens provide clinical phenotype context and does not replace biological confirmation or exclude biomarker testing when clinical concern remains high. **Abbreviations:** CSF, cerebrospinal fluid; FAQ, Functional Activities Questionnaire; MCI, mild cognitive impairment; PET, positron emission tomography; p-tau217, phosphorylated tau 217.

**eFigure 7.**
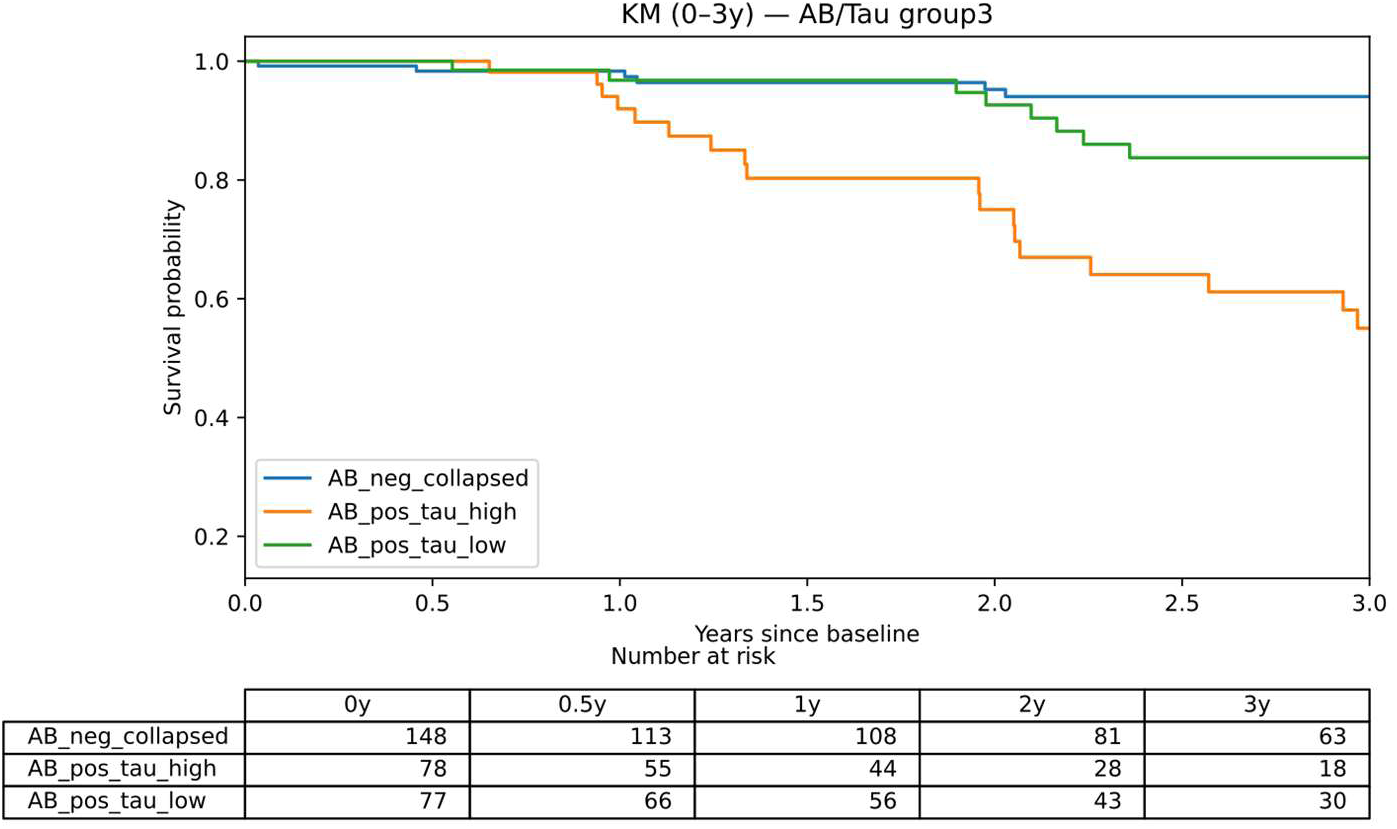
Kaplan-Meier Curves by Amyloid/Tau Biomarker Group. **Note:** Kaplan-Meier curves summarize progression-free probability across collapsed amyloid/tau biomarker groups. The figure provides biological context for the Locked FAQ Trio findings and does not define a primary prognostic comparison. **Abbreviations:** PET, positron emission tomography.

**eFigure 8.**
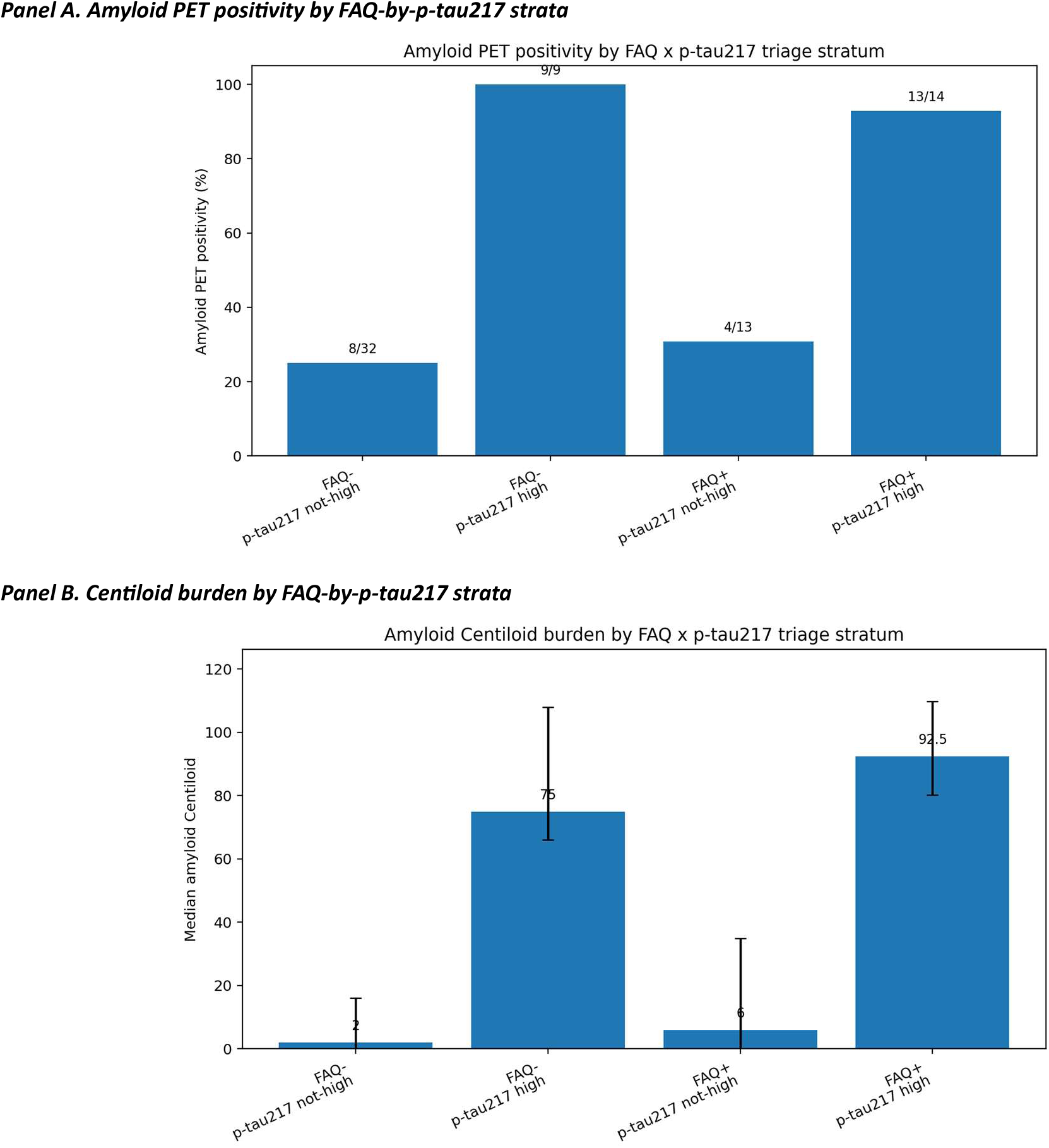
Amyloid PET Positivity and Centiloid Burden by FAQ-by-p-tau217 Strata. **Note:** Panels provide amyloid PET status and continuous amyloid burden context for the descriptive FAQ-by-p-tau217 strata. Component-specific denominators limit interpretation, and these displays do not define a primary prognostic comparison. **Abbreviations:** FAQ, Functional Activities Questionnaire; PET, positron emission tomography; p-tau217, phosphorylated tau 217.

**eFigure 9.**
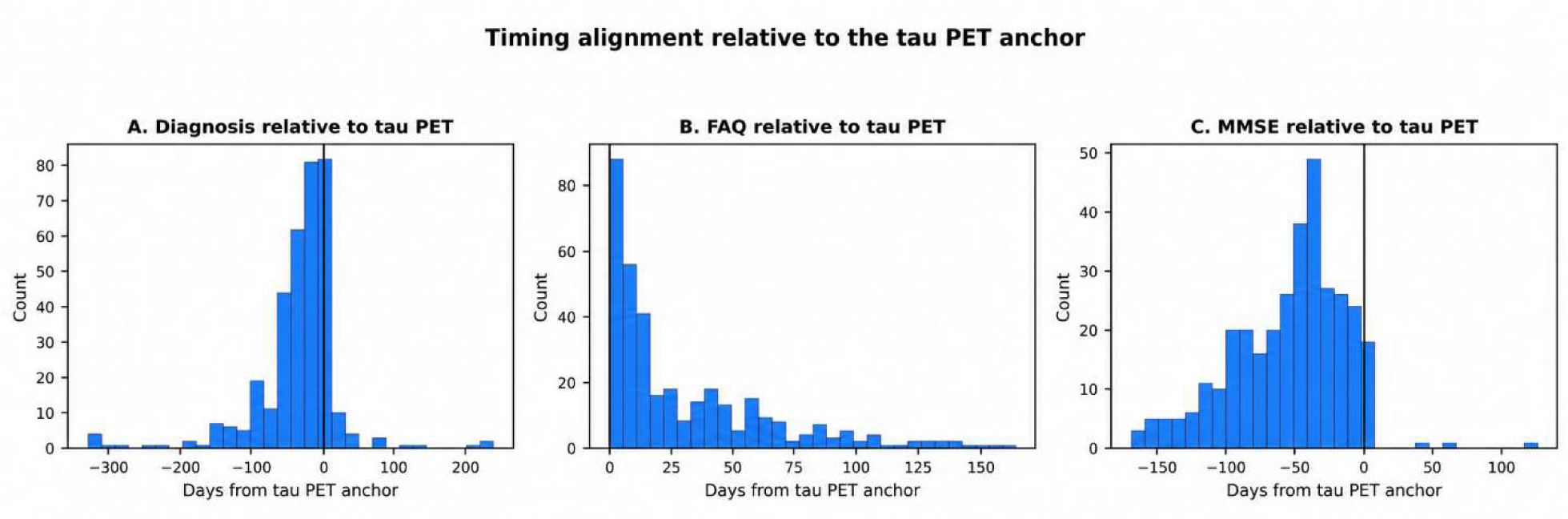
Timing Alignment Relative to the Tau PET Anchor. **Note:** Histograms show timing differences between the tau PET anchor and the nearest diagnostic assessment used to assign MCI status, the matched Functional Activities Questionnaire assessment, and the matched Mini-Mental State Examination assessment. The vertical line indicates the tau PET anchor. Negative values indicate assessments before tau PET, and positive values indicate assessments after tau PET. FAQ assessments were generally close to the tau PET anchor, while MMSE assessments more often preceded tau PET and had a wider timing distribution. This timing structure motivated latest-predictor anchor and 30-day contemporaneity sensitivity analyses. **Abbreviations:** FAQ, Functional Activities Questionnaire; MCI, mild cognitive impairment; MMSE, Mini-Mental State Examination; PET, positron emission tomography.

